# Diarrhea, Helminth Infection, Dehydration, and Malnutrition Associated with Water, Sanitation, and Hygiene Facilities and Poor Handwashing in Schools in Metro Manila, Philippines: A Cross-Sectional Study

**DOI:** 10.1101/2020.12.13.20248141

**Authors:** Stephanie O. Sangalang, Nelissa O. Prado, Allen Lemuel G. Lemence, Mylene G. Cayetano, Jinky Leilanie DP. Lu, John Cedrick Valencia, Thomas Kistemann, Christian Borgemeister

## Abstract

Diarrhea, soil-transmitted helminth (STH) infection, and malnutrition threaten the lives of millions of children globally but particularly in the Global South, where inadequate water, sanitation, and hygiene (WaSH) drive disease risk. The aim of our study was to identify environmental and behavioral risk factors of these diseases among schoolchildren in Metro Manila, Philippines. We analyzed data from a multistage cluster sample of grade 5-10 students to investigate WaSH facilities and hygiene practices. Outcomes were self-reported diarrhea and STH infection and observed malnutrition (stunting, undernutrition, over-nutrition); we used logistic regression models to explore correlates. We included 1,558 students from 15 schools in 3 cities. Over 14% (212) of students experienced diarrhea only, 29.7% (438) experienced STH infection only, and 14% (207) experienced both diarrhea and STH infection. Over 15% (227) of students were stunted, ∼6% (87) were undernourished, and 21% (306) were over-nourished. While diarrhea was associated with poor handwashing, avoiding school restrooms, and lack of a restroom cleaning policy, STH infection was associated with students’ dissatisfaction with school restrooms. Risk of having both diarrhea and STH infection increased when school restrooms lacked water or were unclean. Being only stunted was associated with diarrhea, while being both stunted and undernourished was associated with STH infection. These findings demonstrate that adequate water supply and cleanliness of school WaSH facilities must be achieved and maintained to prevent disease. Future school-based WaSH interventions are recommended to provide clean WaSH facilities that have water, promote handwashing, and discourage avoidance of school toilets.

## Introduction

Diarrhea and soil-transmitted helminth (STH) infection affect 2.39 billion and > 1.9 billion people, respectively.^1, 2^ In 2016 diarrhea caused 1,655,944 deaths.^1^The same year about 60% of diarrhea deaths (829,000) were attributed to inadequate water, sanitation, and hygiene (WaSH).^3^ Nearly 90% of deaths occurred in South Asia and sub-Saharan Africa.^4^ People of low socioeconomic status (SES) are disproportionately affected due to their high exposure to risk factors, e.g. inadequate WaSH facilities and food insecurity. While many on-going programs to prevent diarrhea and STH infection target children < 5, it is uncertain how older children are being affected. School-age children and teenagers should not be ignored as they also bear heavy burdens of diarrheal disease^5^ and STH infection^6, 7^ and may experience high rates of school absenteeism.^8, 9^ STH infection in 2015 and diarrhea in 2016 resulted in ∼6.1 million^2^ and 74.4 million disability-adjusted life-years (DALYs),^1^ respectively. Diarrhea has been associated with high annual costs, ranging from USD $1.3-$1.7 million in Rwanda^10^ to USD $926.4 million in China.^11^

Malnutrition affects ∼1 out of 3 people globally.^12^ In 2017 stunting affected 22.2% (150.8 million) of all children < 5. In 2017, while 5.6% (38.3 million) of children were overweight, 7.5% (50.5 million) were wasted or thin.^13^ Undernutrition, especially in low- and middle-income countries (LMICs), caused 45% of deaths in children < 5.^14^ Overweight and obesity caused ∼7% of deaths (4 million) and 120 million DALYs.^4^ Malnutrition, in all its forms, costs society ∼USD $3.5 trillion, or 5% of the global gross domestic product (GDP), annually.^15^

Preventing children’s malnutrition involves preventing infectious diseases that precipitate imbalanced protein and/or energy intake. Infectious diseases, in turn, can be prevented by improving WaSH, i.e. interrupting routes of fecal-oral disease transmission through proper handwashing, safe handling of food and disposal of feces, and providing access to clean water. In 2016, 6,000 deaths due to malnutrition could have been prevented by improving WaSH.^3^ Community-based WaSH interventions have been associated with decreased risk of diarrhea and STH infection, and consequently, decreased risk of malnutrition.^16, 17^ However, it is unclear how school-based WaSH interventions can benefit children in megacities, or cities with > 10 million inhabitants.^18^

The Philippines, an archipelagic country comprised of 7,641 islands located in Southeast Asia, had a population of ∼106.7 million in 2018.^19^ It has a tropical monsoon climate, with dry (December-May) and wet seasons (June-November), and is located in the Ring of Fire, also known as the Circum-Pacific Belt, which is where 75% of the world’s volcanoes are found and where 90% of all earthquakes take place.^20^ The Philippines’ National Capital Region (NCR), known as Metro Manila (MM), is a megacity that had ∼12.9 million inhabitants in 2015,^21^ comprising 12.1% of the country’s population. In 2015 MM’s population density was 20,785 persons per km^2, 22^ which was > 4× the population density of Beijing in 2014.^23^ MM has a unique risk profile as a megacity that is exposed to > 3 types of natural disasters,^18^ e.g. typhoons, floods, and volcanic eruptions. Thus, MM represents an important intersection of human health and the environment. Due likely in large part to the rapid pace of population growth and impaired institutional capacities in MM, gaps in environmental health management have contributed to the increased prevalence of environment-related infectious diseases such as diarrhea and STH infection. This is an important issue because infectious diseases may trigger or exacerbate malnutrition, which has long-term health consequences, e.g. stunting^1^ and reduced cognitive and verbal performance.^24^

In 2016, 14,800 deaths, including ∼4,113 from diarrhea and ∼48 from STH infection, were attributed to inadequate WaSH in the Philippines.^3^ Diarrhea is the 6^th^ leading cause of disease in MM and among the top 10 causes of disease nationally.^25^ STH infection was endemic in 16 out of 17 Regions in the Philippines, with a prevalence of > 50%.^26, 27^ In spite of de-worming programs, the prevalence rate of STH infection remains high, found to be 66% in preschool-age (12-71 months old) children^28, 29^ and 54% in primary school (grade 3) children.^30^ Prevalence of STH infection in older children was 31.3% in a 2014 survey of 14-15 year-old secondary school students.^31^ In 2018 the prevalence rates of school-age (6-10 years old) children’s stunting, underweight, wasting/thinness (low weight-for-height), and overweight-for-height were: 24.5%, 25%, 7.6%, and 11.7%, respectively.^32^ Over 29,000 annual deaths of children < 5 in the Philippines were attributed to undernutrition.^33^ While undernutrition alone costs the Philippines USD $4.4 million annually,^33^ the overall cost of hunger was USD $6.5 billion in 2013.^34^

The Department of Education (DepEd), Philippines, operates 54,602 public schools nationally and hosted > 22.6 million children during school year (SY) 2018-2019.^35^ While public schools receive government funding, they are severely under-staffed, have a shortage of classrooms, and are overcrowded. The student-to-classroom ratio in public schools in MM ranges from 50:1 to over 100:1.^35, 36^ This has caused the implementation of “double-shift” school days wherein one-half of students attend school during a morning shift (6:00 AM-12:00 PM) and one-half of students attend school during an afternoon shift (12:00 PM-6:00 PM). In MM, the problem has worsened to the point of 7 schools introducing a “triple-shift” in 2019.^37^ The country’s poorest children attend public schools where their vulnerability toward diarrhea and STH infection tends to increase. One reason may be the school environment, which they are regularly exposed to for prolonged periods of time. The purpose of this study was to assess risk factors of diarrhea and STH infection before implementing a school-based WaSH intervention program in MM. We collected data on students’ health and nutrition status, hygiene practices, and WaSH-related perceptions, as well as schools’ WaSH facilities and relevant policies, in order to find out if certain child- or school-level factors increased children’s risk for diarrhea or STH infection. We collected data from a subsample of children’s households, assessing demographic information, families’ handwashing, food security, and homes’ WaSH facilities.

## Materials and Methods

### Study Setting and Design

We conducted a school-based survey on a multistage cluster sample of primary and secondary school students from 15 public schools in MM, where prevalence rates of diarrhea and STH infection are high but access to environmental health and educational programs aimed at disease prevention is low. This paper describes the baseline study that took place during the dry season and the beginning of the wet season (February-June 2017), was observational, and part of a larger research project, “WaSH in Manila Schools”, which involved developing and evaluating a comprehensive, school-based WaSH intervention package. We focused on 2 cities in MM, Navotas and Quezon City (Figure 1), because they are considered geographically and sociodemographically representative of the 14 other cities in MM. Our sampling frame (Figure 2) was the total number of public schools in Navotas and Quezon City, which consisted of 164 schools in 160 “barangays” (the smallest government units in the Philippines) in 8 legislative districts.

**Figure 1.**
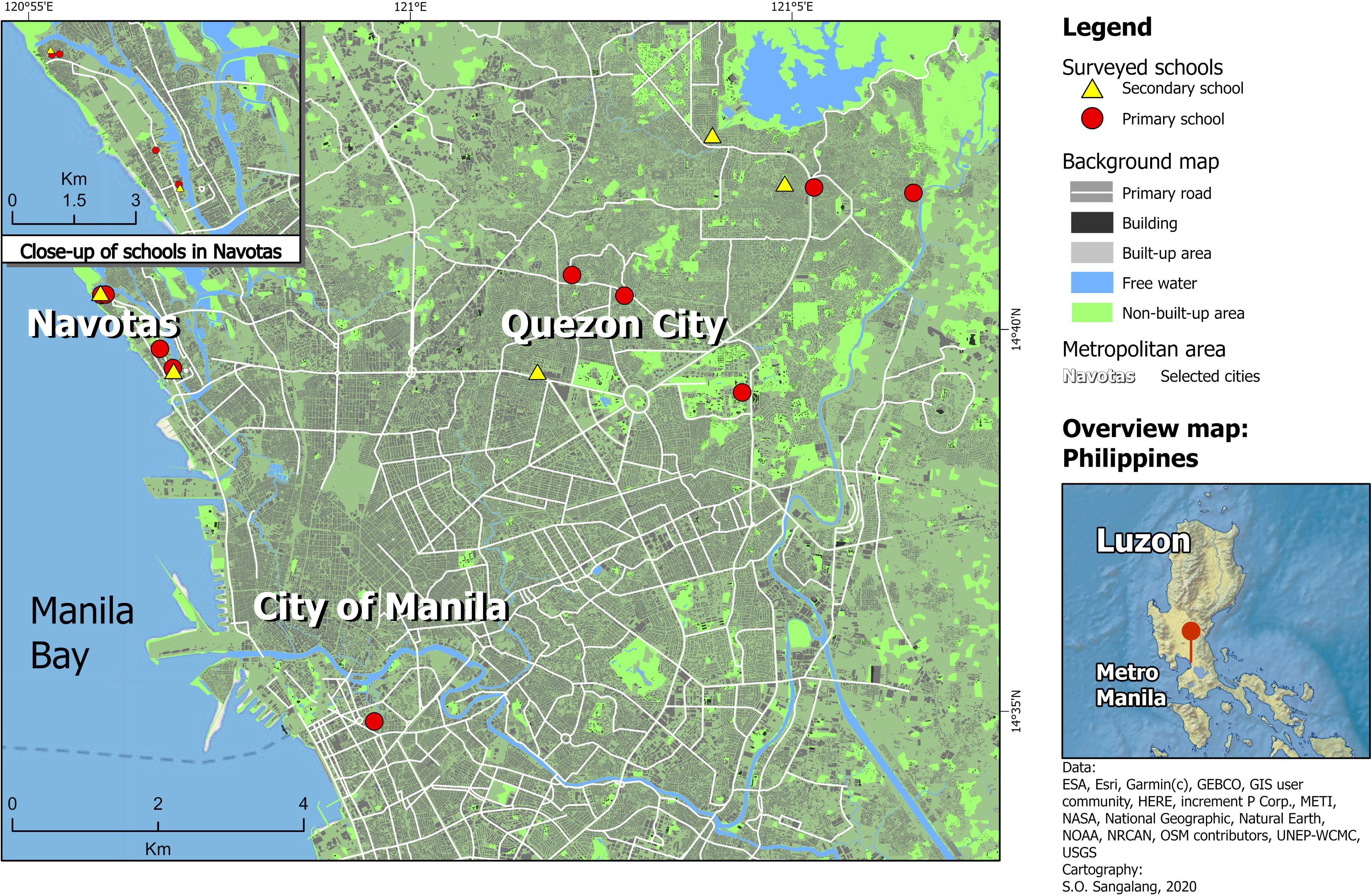
Map of study area with points marking the location of public schools wherein we conducted this observational study during February - June 2017. The map shows the study area in the National Capital Region (NCR), also known as Metro Manila (MM), in northern Philippines, with study schools plotted as points. The red pushpin marker in the lower left inset map indicates where the location of the study area is within the Philippines, specifically in the northern most island group of Luzon.

**Figure 2.**
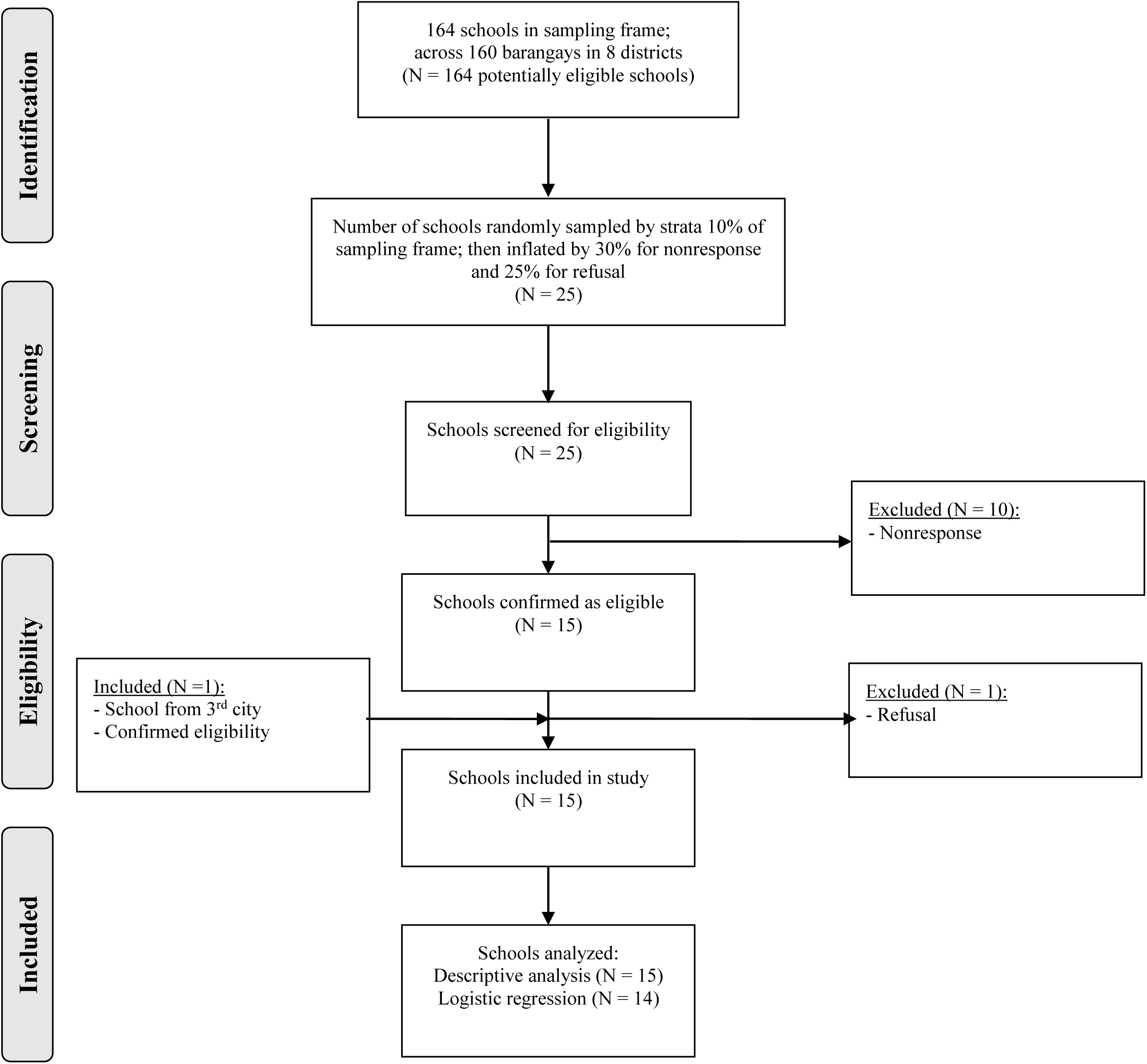
Flow diagram of recruitment of public schools showing school selection, inclusion, and analysis.

During the first stage of sampling we obtained annual school enrollment data for SY 2015-2016 from the DepEd,^35^ identified all public primary schools in Navotas and Quezon City, and sorted them by annual enrollment size, from largest to smallest. We randomly selected 25 schools from the top of the sorted list to invite to participate in our study. After applying our inclusion criteria ([1] accessibility of classrooms during school-day hours; [2] availability of WaSH facilities, e.g. toilets, handwashing basins), we identified 15 schools to visit and ask permission from school principals to conduct our survey. One of the 15 schools had served as the location of our previous pilot study wherein we tested our survey instruments. A 16^th^ school, located in the city of Manila, asked us directly to participate in our study. Before we finished our recruitment of study participants, one of the schools we invited to join our study refused to participate. Thus, our final study sample came from 15 out of 16 originally contacted schools in 3 cities (participation rate: 93.8%). During the second stage of sampling we asked school principals, or representatives who were familiar with students’ schedules, to select the class section(s) that we would survey based on scheduling availability to help us comply with the DepEd’s “no disruption of class” policy. Applying our inclusion criteria, of recruiting students in grade 5 or 6 from primary schools and students in grade 7, 9, or 10 from secondary schools, school principals/representatives selected the class section(s), while keeping the research team blinded. Based on the school’s enrollment size, 1-3 class sections per school were selected in order to obtain a target sample of ∼100 students per school. All students belonging to the selected class section(s) were invited to participate in our study if they met our inclusion criteria: able to 1) read, comprehend, and answer our questionnaire; 2) operate an electronic tablet independently or with minimal assistance; 3) provide a urine specimen; 4) be measured for height and weight.

Prior to starting field research, we estimated the sample size. Our target population was all the public school children in MM. There was a total of 2,059,447 public school children (1,373,852 elementary and 685,595 secondary school children) in MM in SY 2014-2015.^35^ We used the Lynch formula^38^ to estimate the study sample size of ∼384 school children. We inflated the sample by 30% to account for nonresponse and 45% for refusal. To account for differences in schools’ enrollment sizes and the possible effects of the study design, we inflated the sample by another 45% and 20%, respectively. Our target sample size was *N* = 1,308 and 1,558 students enrolled in the study. We received complete responses to questions about our study’s outcomes and exposures of interest from 1,296 students (response rate: 83.2%). We conducted household surveys on a subsample of students and their parents/guardians as described below.

We developed a self-administered questionnaire (Table S1) in English for students and then translated it into Filipino language, also known as Tagalog. We developed structured interview scripts for school principals and parents, school and home restroom inspection checklists, and a students’ health examination data entry form. We pilot tested these electronic survey instruments (available as preprints; please see end of article) at one school, and then refined them to improve understandability. The final versions of the survey instruments were administered using the QuickTapSurvey^©^ app installed on electronic tablets. We preserved the students’ health examination data entry form as a Microsoft Excel^©^ file. Research assistants received hands-on training from the research supervisor during a one-day workshop prior to conducting field research. We went to the selected schools to explain our study’s objectives and procedures to school principals, presented our endorsement letter from the superintendent of the school district, requested the school principal’s written permission to conduct the study, and confirmed the schedule for our survey. We offered school principals no monetary reimbursement.

When we returned to the schools to conduct the survey, research assistants gave a simple explanation in Filipino language about the study to the students, who were assigned a study identification number (ID). This enabled students to provide data anonymously, thereby helping us to ensure confidentiality. Research assistants measured students’ height (without shoes), using a standard tape measure attached to the classroom or hallway wall, and weight (without shoes or items inside their pockets), using a standard digital weighing scale [EKS Asia Ltd., Hong Kong, Special Administrative Region (SAR) of the People’s Republic of China (PRC)]. Research assistants performed point-of-care urinalysis on students’ urine specimens, using urine test strips [Insight Urinalysis Reagent Strips, Acon Laboratories Inc., San Diego, California, U.S.A.]. Research assistants completed the school restroom inspection checklist and took digital photos of WaSH facilities; they also interviewed school principals or representatives about WaSH policies. The research supervisor verified adherence to research protocols via direct observation. We obtained other school data, e.g. number of classrooms, annual budget for Maintenance and Other Operating Expenses (MOOE), from the DepEd.^35, 36, 39^

### Ethics

The ethics committees at the University of Bonn, Germany (Number 216/16), and the University of the Philippines (U.P.) Manila (Number 2017-0113), gave written approval for the study. Before conducting the school surveys, we obtained written approval from local authorities, i.e. superintendents of school districts representing the DepEd. Written informed consent was provided by school principals in *loco parentis* (translation: “in the place of a parent”) for the students’ participation. We explained in Filipino language the study objectives and procedures to the teachers and students, and said that participation in the study was voluntary and results would remain confidential and have no impact on students’ school grades.

### Outcome Definitions

We measured the primary outcomes of diarrhea and STH infection prevalence via students’ self-report assessed by a questionnaire, which included specific binary (yes no) questions. The rationale for using self-report to assess diarrhea prevalence were: reliability, validity, convenience, and the ability to quickly, affordably, and accurately assess prevalence in a large sample of children. Self-reported diarrhea has been used in previous studies involving schoolchildren in LMICs.^40–42^ In our study, diarrhea was defined as a “yes” answer to one question in the questionnaire (“Did you have diarrhea in the past month?”). We used the WHO’s case definition of diarrhea, i.e. having >= 3 loose/liquid bowel movements (i.e. stools) in one day.^43^ We estimated diarrhea prevalence by dividing the number of children who answered “yes” by the total number of children who answered the question, either “yes” or “no”. We then multiplied the output by 100 to obtain the prevalence rate.

STH infections are often assessed via diagnostic tests such as the Kato-Katz technique, which involves counting the number of eggs per gram of feces. In our study, examining children’s feces would not have been feasible due to financial, time, and personnel constraints. Furthermore, it would not have been appropriate or necessary for two reasons: 1) we aimed to measure STH infection prevalence, not STH infection intensity (thus, an egg count was not needed); 2) in the Philippines, public school children living in STH-endemic areas, such as out study site, Metro Manila, undergo biannual deworming. Thus, a child who recently underwent deworming would likely test negative for STH infection (no eggs or worms would be found in his/her feces). This would have resulted in a falsely lower STH prevalence rate, which failed to accurately measure the burden of disease. Another disadvantage of using feces-based measures was that some children may have felt too embarrassed to provide us with a specimen of their feces. For these reasons, we decided to use children’s self-report as the measure for STH infection. The rationale for using self-report were: accuracy (depending on children’s recall ability), convenience, and affordability. Self-report enabled us to quickly and easily assess STH infection prevalence in a large study sample. Evidence is limited about using self-report to assess STH infection. However, evidence is available about using children’s self-report to assess STH infection-related knowledge, attitudes, and practices (KAP).^44, 45^ In our study, we defined STH infection as a “yes” answer to one question in the questionnaire (“Have you ever had ‘worms’?”). We estimated STH infection prevalence by using a proportion: a) we counted the number of children who answered “yes” to the above question; b) we counted the total number of children who answered “yes”, “no”, and “I don’t know” to the same question; we calculated the prevalence rate by dividing a) by b) and multiplying the output by 100.

We measured the secondary outcomes of stunting, undernutrition, and over-nutrition as follows: 1) we used anthropometry to measure children’s height and weight; 2) we used the WHO AnthroPlus software (WHO, Geneva, Switzerland) to calculate z-scores; 3) we used the WHO 2007 Growth Reference for children 5 - 19 years old^46^ and WHO’s cut-off points to classify children as stunted and under-nourished or over-nourished.^47, 48^ “Stunting” was defined as having a height-for-age z-score (HAZ) < -2. We considered “undernourished” to be a composite variable, i.e. comprised of two variables. First, we considered “underweight” (or “thin”), which is based on body mass index (BMI)-for-age z-scores (BAZ). BMI is calculated with the formula: weight (kg) / [height (m)]^2^. The cut-off points of z-scores for “underweight” are -3 < BAZ < -2. Second, we considered “wasted” (or “severely thin”), which is also based on BAZ. The cut-off point of z-scores for “wasted” is BAZ < -3.

The rationale for using BAZ instead of weight-for-height because we wished to capture changes in the weight-height relationship that take place with aging. BAZ may be used to assess individuals continuously up to age 20; this was important for us because our study sample included some older teenagers nearing the age of 20. We did not use weight-for-age because it is recommended for only for children 10 years old and younger. Another disadvantage of weight-for-age is that it does not differentiate between height and body mass during puberty when some children have growth spurts and could appear to have excess weight but actually are just growing taller.^49^ BMI, which is measured in BAZ but not in weight-for-age or weight-for-height, is recommended to be used especially for adolescents.^50–52^

We considered “over-nourished” to be a composite variable, i.e. comprised of two variables. First, we considered “overweight”, which is based on BAZ. The cut-off points of z-scores for “overweight” are 1 < BAZ < 2. Second, we considered “obese”, which is also based on BAZ. The cut-off point of z-scores for “obese” is BAZ > 2.

Acute dehydration was defined as having highly concentrated urine, i.e. urine specific gravity (U_sg_) >= 1.020,^53^ (Table 1), measured using (reagent) urine test strips (Insight Urinalysis Reagent Strips, Acon Laboratories Inc., San Diego, California, U.S.A.). We used the cut-off of U_sg_ 1.020 because it corresponds to a urine osmolality (U_osm_), considered to be the gold standard urine-based measure of dehydration,^54, 55^ of 800 mOsm/kg H2O, which was the cut-off used in previous studies involving dehydrated children.^56, 57^ Urine test strips are a low-cost, convenient way to perform urinalysis in field settings.^58^ The procedure was as follows: 1) we asked all children to provide a urine specimen in a plastic cup labeled with their study ID number; 2) we dipped one urine test strip into one urine specimen; 3) after waiting for a minimum of 2 minutes, we determined the U_sg_ by matching the color change that appeared on the urine test strip with the manufacturer’s interpretation guide. We assessed all study participants for acute dehydration in this way.

**Table 1.** Operational definitions.

| Indicator |  | Definition | Source |
| --- | --- | --- | --- |
| Malnutrition |  |  |  |
| Stunting | Stunted | HAZ<sup>a</sup> < -2 | 47, 61, 62 |
| Undernutrition | Underweight | -3 < BAZ<sup>a,b</sup> < -2 | 47, 59, 60 |
|  | Wasted (“severely thin”) | BAZ < -3 | 47, 59, 60 |
| Over-nutrition | Overweight | 1 < BAZ < 2 | 47, 59, 60 |
|  | Obese | BAZ > 2 | 47, 59, 60 |
| WaSH adequacy |  |  |  |
| Sanitation |  |  |  |
| Improved | Facility that hygienically separates human excreta from human contact | e.g. flush toilet, pour-flush latrines, ventilated improved pit latrines and pit latrines with a slab or covered pit | 143 |
| Unimproved | Facility that does not hygienically separate human excreta from human contact | e.g. pit latrines without slabs or platforms or open pit, hanging latrines, bucket latrines, open defecation, disposal of human feces with other forms of solid waste | 143 |
| Male student-to-toilet ratio | Less than 50 students | 1 toilet, 1 urinal, 1 handwashing basin | 63 |
|  | 50 or more students | 2 toilets, 1 urinal, 2 handwashing basins | 63 |
|  | For each additional 100 students | 1 toilet, 1 urinal, 1 handwashing basin | 63 |
| Female student-to-toilet ratio | Less than 30 students | 1 toilet, 1 handwashing basin | 63 |
|  | 30 - 100 students | 2 toilets, 2 handwashing basins | 63 |
|  | For each additional 50 students | 1 toilet | 63 |
|  | For each additional 100 students | 1 handwashing basin | 63 |
| Male student-to-toilet ratio | 50 students | 1 toilet, 1 urinal (or 50 cm of urinal wall) | 64 |
| Female student-to-toilet ratio | 25 students | 1 toilet | 64 |
| Student-to-handwashing (hw) basin ratio | N/A | N/A | No WHO guidelines available |
| Male student-to-toilet ratio | Low | ≤ 50:1 | Category used in present study |
|  | Medium | 51:1–100:1 | Category used in present study |
|  | High | ≥ 101:1 | Category used in present study |
| Female student-to-toilet ratio | Low | ≤ 50:1 | Category used in present study |
|  | Medium | 51:1–100:1 | Category used in present study |
|  | High | ≥ 101:1 | Category used in present study |
| Student-to-hw basin ratio | Low | ≤ 50:1 | Category used in present study |
|  | Medium | 51:1–150:1 | Category used in present study |
|  | High | ≥ 151:1 | Category used in present study |

Note: BAZ, body mass index-for-age Z-score; DOH, Department of Health, Philippines; HAZ, height-for-age Z-score; hw, handwashing; N/A, not applicable; WaSH, water, sanitation, and hygiene; WHO, World Health Organization.
<sup>a</sup>HAZ and BAZ estimates were obtained using WHO AnthroPlus software (WHO, Geneva, Switzerland). Estimates are based on the WHO Reference 2007 for children 5-19 years old.
<sup>b</sup>BAZ based on body mass index (BMI), which is calculated by weight (kg) / [height (m)]<sup>2</sup>.

Malnutrition was defined according to the WHO guidelines (Table 1). We considered “undernourished” to be a composite variable, i.e. comprised of two variables. First, we considered “underweight” (or “thin”), which is based on body mass index-for-age Z-scores (BAZ). BAZ is based on height and weight, using the following formula: weight (kg) / [height (m)]^2^. The cut-off points of Z-scores for “underweight” are -3 < BAZ < -2. Second, we considered “wasted” (or “severely thin”), which is also based on BAZ. The cut-off point of Z-scores for “wasted” is BAZ < -3. We considered “over-nourished” to be a composite variable, i.e. comprised of two variables. First, we considered “overweight”, which is based on BAZ. The cut-off points of Z-scores for “overweight” are 1 < BAZ < 2. Second, we considered “obese”, which is also based on BAZ. The cut-off point of Z-scores for “obese” is BAZ > 2. BAZ estimates were obtained using WHO AnthroPlus software (WHO, Geneva, Switzerland). BAZ estimates were based on the WHO Reference 2007 for children 5-19 years old.^47, 59–62^ Cut-off points of Z-scores were from the WHO’s Growth Reference 5 - 19 years, BMI-for-age (5 - 19 years).^48^

### Exposure Definitions

We assessed risk factors at the individual- and school-levels. We defined children as individuals < 13 years old and teenagers as individuals >= 13 years old. We asked students about handwashing, use and perceptions about school WaSH facilities, health history, nutrition, and if hygiene lessons were taught in school. We asked school principals about WaSH-related school policies, e.g. “Is there a school policy to clean the students’ restrooms daily?”. We counted the number and assessed the quality of school WaSH facilities, noting characteristics of improved versus unimproved sanitation (Table 1).

We estimated student-to-toilet and student-to-handwashing basin ratios based on the Department of Health (DOH), Philippines, guidelines^63^ (Table 1). The DOH guidelines do not include specific or fixed ratios, rather they recommends a range of numbers for WaSH facilities that are sex-specific. For “50 or more” male students, a student-to-toilet ratio of 50:2 and a student-to-handwashing basin ratio of 50:2 are recommended; for each additional 100 male students, one toilet and one handwashing basin are recommended.^63^ For 30 - 100 female students, two toilets and two handwashing basins are recommended; for each additional 100 female students, one toilet and one handwashing basin are recommended.^63^ We decided not to base our estimations on the WHO guidelines^64^ (Table 1) because public schools in many parts of the Philippines, similar to other countries in the Global South, currently have limited capacity to effectively address the over-crowding of students on school campuses.

We conducted a survey with a subsample of students’ households to assess risk factors, e.g. food security, access to drinking water, at the home-level. If a student provided a functioning telephone number during the questionnaire portion of the school survey, then his/her parent/guardian was contacted by a research assistant to be recruited for the household survey. Our target sample of parents/guardians was 10-12 per school. To account for nonresponse or refusal, we inflated the sample by 10%. Thus, our target sample size for the subsample was *N =* 225. Research assistants explained over the telephone our study’s objectives and procedures, and then asked for permission from parents/guardians to visit the home on a later date. Research assistants worked in pairs to conduct the household survey in person: one research assistant interviewed the parent/guardian, while the other research assistant inspected the home’s restroom.

We collected samples for water quality testing from study schools and a separate sample of households (located in the school neighborhood) in April 2018, after the post-intervention assessment of the larger research project, “WaSH in Manila Schools”. We report our water quality indicators in Table S2; we assessed these according to the Philippine National Standards for Drinking Water of 2017.^65^ No health-related guideline values from the Philippines or the WHO^66^ were available for two parameters (biological oxygen demand [BOD] and dissolved oxygen [DO]). Thus, we used guidelines from the United States Environmental Protection Agency (EPA).^67^ For DO, the recommended values were estimated by using a EPA-provided reference table; estimations were dependent on temperature and salinity.

### Statistical Methods

We downloaded data from the QuickTapSurvey^©^ app as Microsoft Excel^©^ files. We used key matching data (students’ self-reported date of birth and telephone number) to link data from students’ questionnaires and health examinations to home restroom inspections and parent/guardians’ interviews. We used Stata, version 15 (StataCorp, College Station, Texas, U.S.A.), to prepare data for analysis.

To describe exposure to inadequate WaSH, we measured frequencies and interquartile ranges (IQRs) relevant to schools’ and homes’ WaSH facilities. Data from school inspections were summarized at the school-level by measuring the mean scores of individual facility inspections. To describe outcomes of diarrhea, STH infection, and malnutrition, we measured prevalence rates of diseases using contingency tables with estimates of standard error (SE) and 95% confidence intervals (CI).

In this study, we used multiple logistic regression, which produces adjusted odds ratios (AORs) rather than non-adjusted odds ratios (ORs). This means that all variables included in the multiple logistic regression model were adjusted for confounding. All results presented in this paper are the outcome of multiple logistic regression analysis.

To identify statistically significant factors associated with diarrhea, STH infection, and malnutrition, we used multiple logistic regression (Box S1), which produces adjusted odds ratios (aORs). In contrast to unadjusted (“crude”) ORs, aORs allowed us to control for confounding. We considered the following potential confounders: children’s sex, age group, malnutrition status, hygiene behaviors, and WaSH-related perceptions, and schools’ WaSH facilities (quantity, quality) and related policies and budget. All results presented in this paper are the outcome of multiple logistic regression analysis. We considered p-values < 0.05 to be statistically significant.

For all multiple regression models, clustering was controlled for using the “cluster” option in STATA. It is important to account for clustering because of the potential for within-group (“intragroup”) correlation among children from the same school and adjust the SE of estimates. The “cluster” option in STATA enabled us to indicate that the observations were clustered into schools (based on school ID) and that the observations may be correlated within schools, but would be independent between schools.

We analyzed the main study sample with 2 multiple logistic regression models: model A for diarrhea only, STH infection only, and dual infection (both diarrhea and STH infection), and model B for malnutrition, i.e. stunting only, underweight only, and over-nutrition only. We included variables such as student does not wash hands in school, school restroom lacks water, and lack of policy to clean school restroom daily. We analyzed the subsample with 2 multiple logistic regression models: model C for diarrhea only, STH infection only, and dual infection (both diarrhea and STH infection), and model D for malnutrition, i.e. stunting only, underweight only, and over-nutrition only. We included variables such as the home restroom is not clean, it has signs of mold, and the number of adults in the home.

## Results

### Study Population

We measured diarrhea, STH infection, and malnutrition prevalence in 1,558 students from 15 schools in 3 cities (Table 2). Students were 9-19 years old; 66.7% (1,039) were < 13 years old and 73.1% (1,085) considered themselves to be “healthy” (Table 3). Over 16% (239) of students said they avoided the school restroom and while > 91% of students (1,359) said they washed their hands at school, only 53% (554) said they washed their hands with soap and water at school.

**Table 2.** Schools included in survey by “barangay”, the smallest government unit in the Philippines (*N* = 15).

| Barangay | Schools (%) | Students (%) |
| --- | --- | --- |
| Location near/adjacent to |  |  |
| River/estuary |  |  |
| Paco | 1 (6.7) | 111 (7.1) |
| Bagong Silangan, North Fairview | 2 (13.3) | 234 (15.0) |
| Coastline/bay |  |  |
| San Jose, Sipac-Almacen | 3 (20.0) | 303 (19.5) |
| Tangos North | 3 (20.0) | 335 (21.5) |
| Highway/primary road |  |  |
| Commonwealth | 2 (13.3) | 203 (13.0) |
| Pasong Tamo, Tandang Sora | 2 (13.3) | 188 (12.1) |
| Santo Cristo | 1 (6.7) | 90 (5.8) |
| U.P. (Diliman) Campus | 1 (6.7) | 94 (6.0) |
| All areas | 15 (100) | 1,558 (100) |

Note: U.P., University of the Philippines.

**Table 3.** Main sample descriptives of demographic, health, and hygiene-related factors of students (*N* = 1,558) and WaSH-related structural factors of schools (*N* = 15).

|  | n | % (95% CI) |
| --- | --- | --- |
| Student factors (N=1588) |  |  |
| Sex (self-reported) |  |  |
| Female (self-reported) | 861 | 55.3 (52.8, 57.7) |
| School attendance (self-reported) |  |  |
| Attended school in the last 6 months | 1391 | 93.5 (92.2, 94.7) |
| Missed class last year due to "health problem" | 610 | 41(38.5, 43.5) |
| Grade group (observed) |  |  |
| Primary (grade 5 - 6) | 1012 | 65 (62.6, 67.3) |
| Age (self-reported) |  |  |
| Median (IQR) |  | 12 (11, 13) |
| Age group (self-reported) |  |  |
| Child (age < 13 years) | 1039 | 66.7 (64.3, 69.0) |
| Adolescent (age => 13 years) | 518 | 33.2 (30.9, 35.6) |
| Health-related knowledge, perceptions, and hygiene practices (self-reported) |  |  |
| Does not wash hands at school | 129 | 8.67 (7.2, 10.1) |
| Does not use soap when washing hands at school | 491 | 47 (44.0, 50.0) |
| Does not use school restroom | 107 | 7.19 (5.9, 8.5) |
| Avoids school restroom | 239 | 16.1 (14.2, 18) |
| Does not know if he/she had STH infection | 318 | 21.4 (19.4, 23.5) |
| Considers oneself to be "not healthy" | 399 | 26.9 (24.6, 29.1) |
| No provision of hygiene lessons at school | 135 | 9.1 (7.6, 10.6) |
| Infectious disease (self-reported) |  |  |
| Had diarrhea (in the last month) | 421 | 28.5 (26.2, 30.9) |
| Had STH infection (ever) | 647 | 43.6 (41.1, 46.2) |
| Malnutrition<sup>a</sup> (observed) |  |  |
| Stunted | 227 | 15.2 (13.4, 17.1) |
| Undernutrition | 127 | 8.6 (7.2, 10.1) |
| Severely thin | 28 | 1.9 (1.3, 2.7) |
| Thin | 99 | 6.7 (5.5, 8.1) |
| Over-nutrition | 321 | 21.7 (19.6, 23.9) |
| Overweight | 226 | 15.3 (13.5, 17.2) |
| Obese | 95 | 6.4 (5.2, 7.8) |
| Sign of acute dehydration (observed) |  |  |
| Highly concentrated urine, sg $\geq 1.020$ | 956 | 68.0 (65.5, 70.5) |
| School factors |  |  |
| Number of schools with unimproved<sup>b</sup> sanitation present | 1 | 7.1 (0.2-33.9) |
| Number of schools without handwashing (hw) basin | 1 | 14.3 (1.8, 42.8) |
| Type of toilet, latrine (e.g. dry, non-flush) | 1 | 1.3 (0.03, 6.9) |
| Type of toilet, pour-flush | 56 | 74.7 (59.6, 80.6) |
| Type of toilet, flush | 16 | 24.1 (12.0, 30.8) |
| Number of toilet bowls, median (IQR) |  | 17 (5.9, 26.5) |
| Number of hw basins, median (IQR) |  | 5 (4, 12.5) |
| Number of schools that exceeded guidelines for student-to-toilet ratio<sup>c</sup> | 13 | 92.9 (66.2, 99.8) |
| Number of schools that exceeded guidelines for student-to-hw unit ratio<sup>c,d</sup> | 13 | 92.9 (66.2, 99.9) |
| Student-to-toilet ratio, median (IQR) |  | 302.3 (219, 418) |
| Student-to-hw basin ratio, median (IQR) |  | 562 (380.3, 935.1) |
| Number of toilets that had no nearby hw basin | 24 | 30.4 (20.5, 41.8) |
| Number of hw basins not near toilet | 61 | 49.2 (40.1, 58.3) |
| Provision of no separate toilet for females | 8 | 3.6 (1.5, 6.8) |
| No water available | 26 | 32.9 (22.7, 44.4) |
| No soap available | 65 | 82.3 (72.1, 90.0) |

Note: CI, confidence interval; hw, handwashing; IQR, interquartile range; sg, specific gravity; STH, soil-transmitted helminth; WaSH, water, sanitation, and hygiene.
<sup>a</sup>Malnutrition indices are defined in Table 1 and were estimated using WHO AnthroPlus software. We classified malnutrition status according to WHO guidelines.<sup>59-62</sup>
<sup>b</sup>Unimproved sanitation does not hygienically separate human excreta from human contact (Table 1).
<sup>c</sup>We referred to the national guidelines of the Department of Health (DOH), Philippines. We estimated student-to-toilet and student-to-handwashing unit ratios by using school enrollment data from school year (SY) 2016 - 2017. Then we divided the number of students by two to take into account schools' use of a "double shift". We divided the number of students by the total number of toilets, taking into account that some toilets were coed toilets (i.e. used by both males and females). We report sex-specific ratios in Supplemental Materials.
<sup>d</sup>We included in the analysis one school which had no functioning hw stations or basins (i.e. sinks); rather, a hose connected to water was provided for children to wash their hands.

During our assessment of associated risk factors of diseases, we excluded 266 students (17.1%): 167 because of nonresponse on outcomes or logistic regression model covariates and 99 (6.4%) because they attended a school where the school principal declined our request to inspect the school restrooms during the baseline study. (The school principal permitted us to inspect the school restrooms during the subsequent pre-intervention assessment [June-July 2017]; we will report findings in a forthcoming paper.)

#### Subsample

From our main study sample, we found *N* = 211 students whose parent/guardian was willing to participate in our household survey. The subsample of students was ∼2/3 (134) female and ∼73% of students were < 13 years old (Table 4). Households had a median of 6 people (interquartile range [IQR] 4, 8) and the median duration of residence in the home was 13 years (IQR 5, 25). We report results of water quality testing in homes in Table S2.

**Table 4.** Subsample descriptives of demographic, hygiene- and food security-related factors of households and WaSH-related structural factors of homes (*n* = 211).

| Factors | *n* | % (95% CI) |
| --- | --- | --- |
| Student level |  |  |
| Female | 134 | 63.5 (56.6, 70.0) |
| Age < 13 years old | 153 | 72.5 (66.0, 78.4) |
| Home level |  |  |
| Household demographics |  |  |
| Number of people in home, median (IQR) |  | 6 (4, 8) |
| Number of adults in home, median (IQR) |  | 3 (2, 5) |
| Number of kids in home, median (IQR) |  | 3 (2, 4) |
| Duration (years) of residence in home, median (IQR) |  | 13 (5, 25) |
| Household WaSH |  |  |
| Has no restroom inside home | 20 | 10.4 (5.9, 14.3) |
| Has no toilet | 3 | 1.4 (0.3, 4.5) |
| Has no handwashing (hw) basin | 66 | 34.4 (27.7, 41.6) |
| Home restroom has no water available | 8 | 4.2 (1.8, 8.0) |
| Home restroom has no soap available | 25 | 13.0 (8.6, 18.6) |
| Home restroom has no hand towels | 129 | 67.2 (60.1, 73.8) |
| Home restroom has no toilet paper | 187 | 97.4 (94.0, 99.1) |
| Home restroom not clean | 76 | 39.6 (32.6, 46.9) |
| Home restroom has signs of mold | 98 | 51.0 (43.7, 58.3) |
| Home restroom has signs of damage | 139 | 72.4 (65.5, 78.6) |
| Home restroom is not well-lit | 83 | 43.2 (36.1, 50.6) |
| Home restroom has wet floor | 145 | 75.5 (68.8, 81.4) |
| Home restroom door has no lock | 85 | 44.3 (37.1, 51.6) |
| Home restroom has unimproved<sup>a</sup> sanitation | 39 | 20.3 (14.9, 26.7) |
| Home restroom toilet cannot be flushed | 140 | 72.9 (66.0, 79.1) |
| Home restroom has septic tank | 57 | 29.7 (23.3, 36.7) |
| Household food insecurity |  |  |
| Insufficient amount of food at home | 14 | 6.6 (3.7, 10.9) |
| No place nearby to buy food | 6 | 2.8 (1.1, 6.1) |
| Food prices are not affordable | 16 | 7.6 (4.34, 12.0) |
| Not able to buy food | 113 | 53.6 (46.6, 60.4) |
| Has experienced asking/begging someone for food | 165 | 78.2 (72.0, 83.6) |
| Does not eat a variety<sup>b</sup> of food | 13 | 6.2 (3.3, 10.3) |
| Cooks food less often than buys prepared food | 20 | 9.5 (5.9, 14.3) |
| No access to drinking water | 4 | 1.9 (0.5, 4.8) |

Note: CI, confidence interval; hw, handwashing; IQR, interquartile range; WaSH, water, sanitation, and hygiene.
<sup>a</sup>Unimproved sanitation does not hygienically separate human excreta from human contact (Table 1).
<sup>b</sup>Variety of food refers to e.g. fruits, vegetables, meat, and fish.

### Outcome and Exposure Measurements

We found handwashing basins in ∼86% of schools (12); ∼1/3 (26) of handwashing basins lacked water and > 82% (65) lacked soap (Table. 3). Over 1/3 (4) of schools had water that was contaminated by coliform bacteria, while 1/4 (3) had water that was contaminated by *Escherichia coli* (*E. coli*) (Table S2).

Over 1/4 of students (28.5%, 421) reported having diarrhea in the last month and ∼44% (647) reported ever having a STH infection (Table 3). Over 15% (227) of students were stunted, ∼9% (127) had undernutrition, and > 21% (321) had over-nutrition (Table 3). Over 2/3 of students (956) had highly concentrated urine (U_sg_ >= 1.020), indicative of dehydration (Table 3). A greater proportion of males (68.7%, 432) compared to females (67.5%, 524), and a greater proportion of teenagers (72.1%, 354) compared to children (65.9%, 602), had acute dehydration as exhibited by U_sg_ >= 1.020 (Table S3).

### Associations between Diarrhea and STH Infection with School WaSH

In multiple logistic regression models, students’ not washing their hands in school was significantly associated with increased odds (95% CI) of diarrhea only, 1.74 (95% CI: 1.14, 2.68) (Table 5). Students’ avoiding to use the school restroom, 1.66 (95% CI: 1.12, 2.47) and schools’ lack of policy for cleaning the restroom daily, 1.56 (95% CI: 1.31, 1.87) (Table 5) were significantly associated with increased odds of diarrhea only, while schools’ having the maximum MOOE budget was significantly associated with decreased the odds, 0.43 (95% CI: 0.26, 0.70).

**Table 5.**
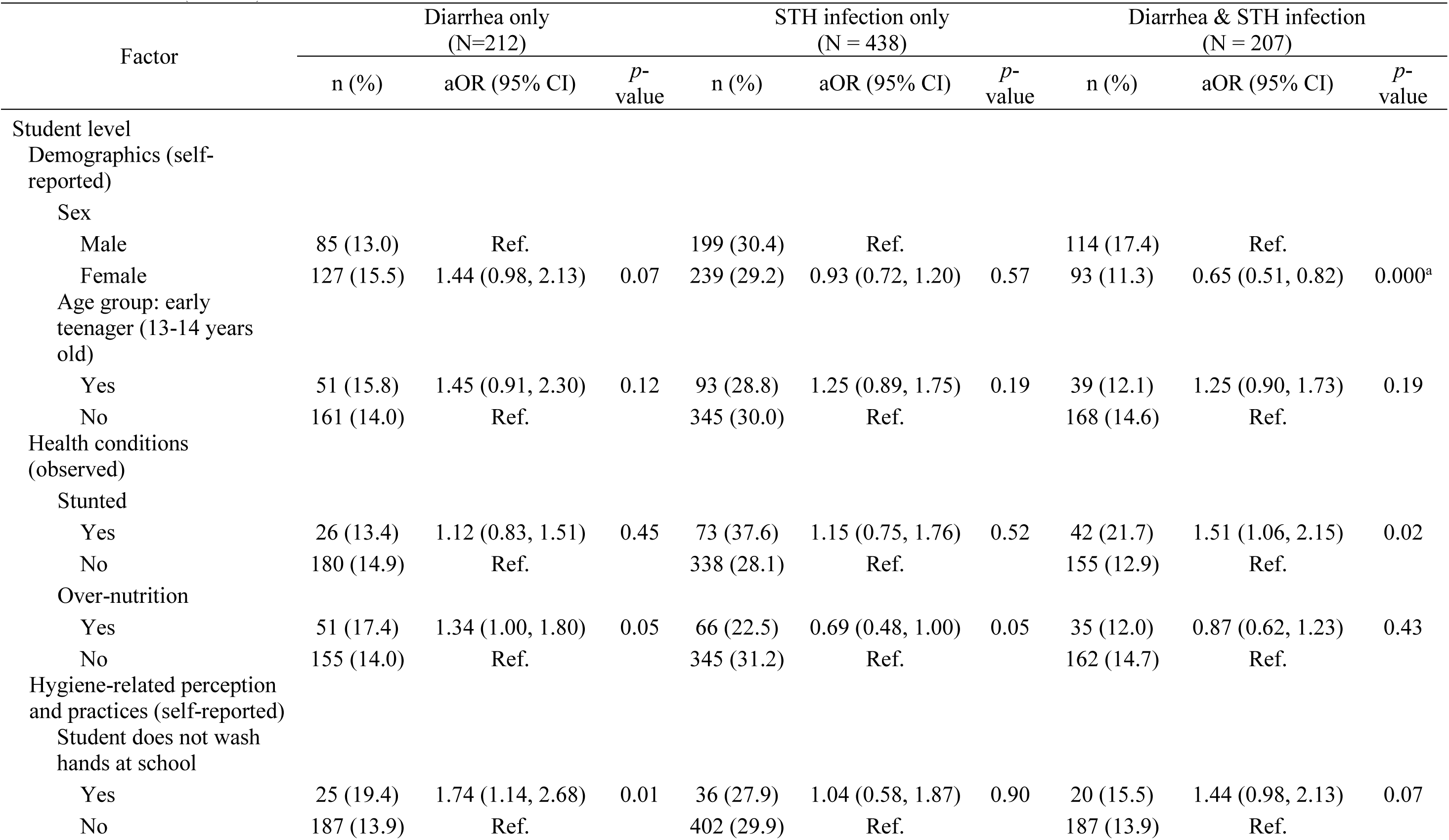
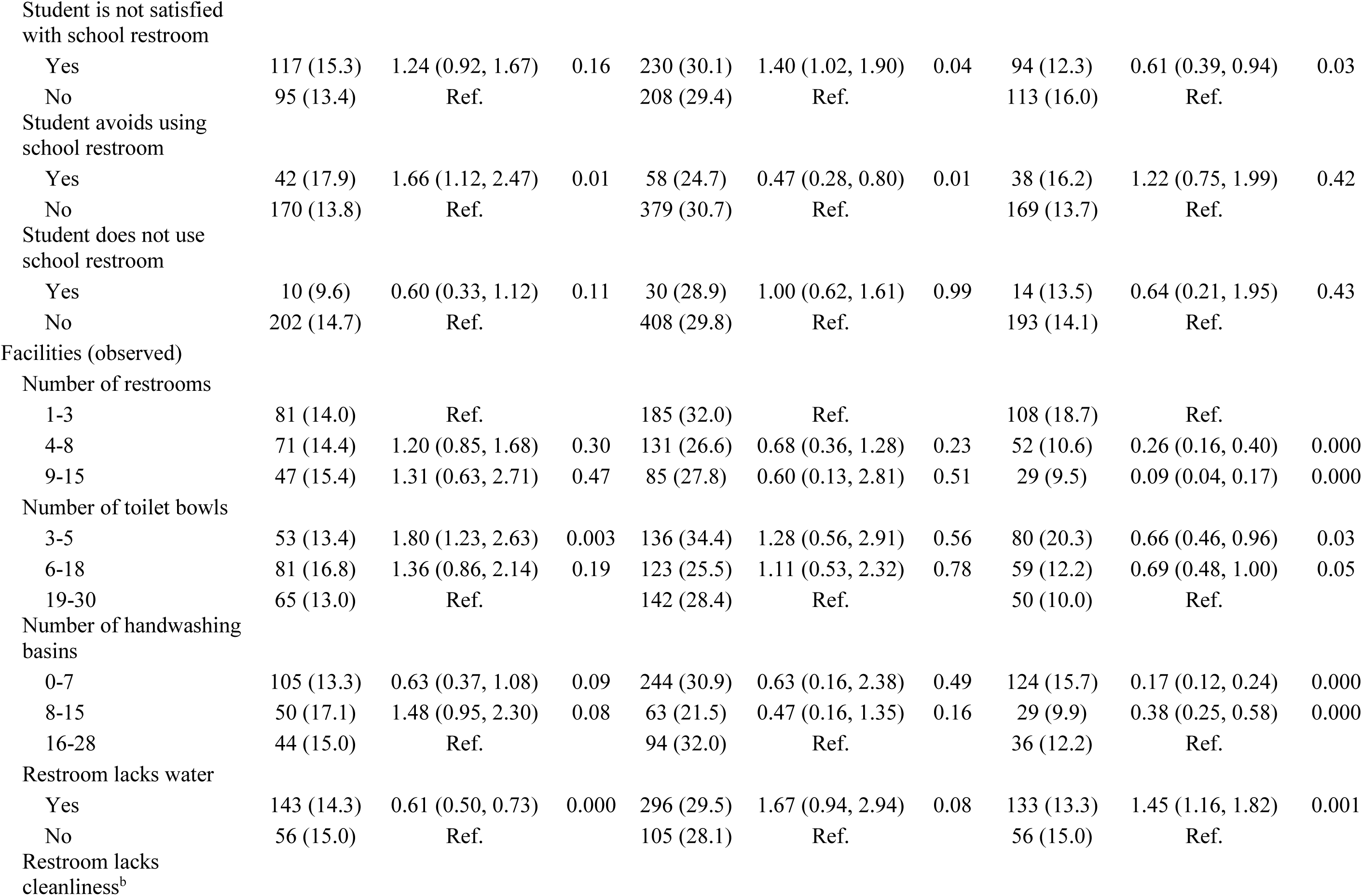
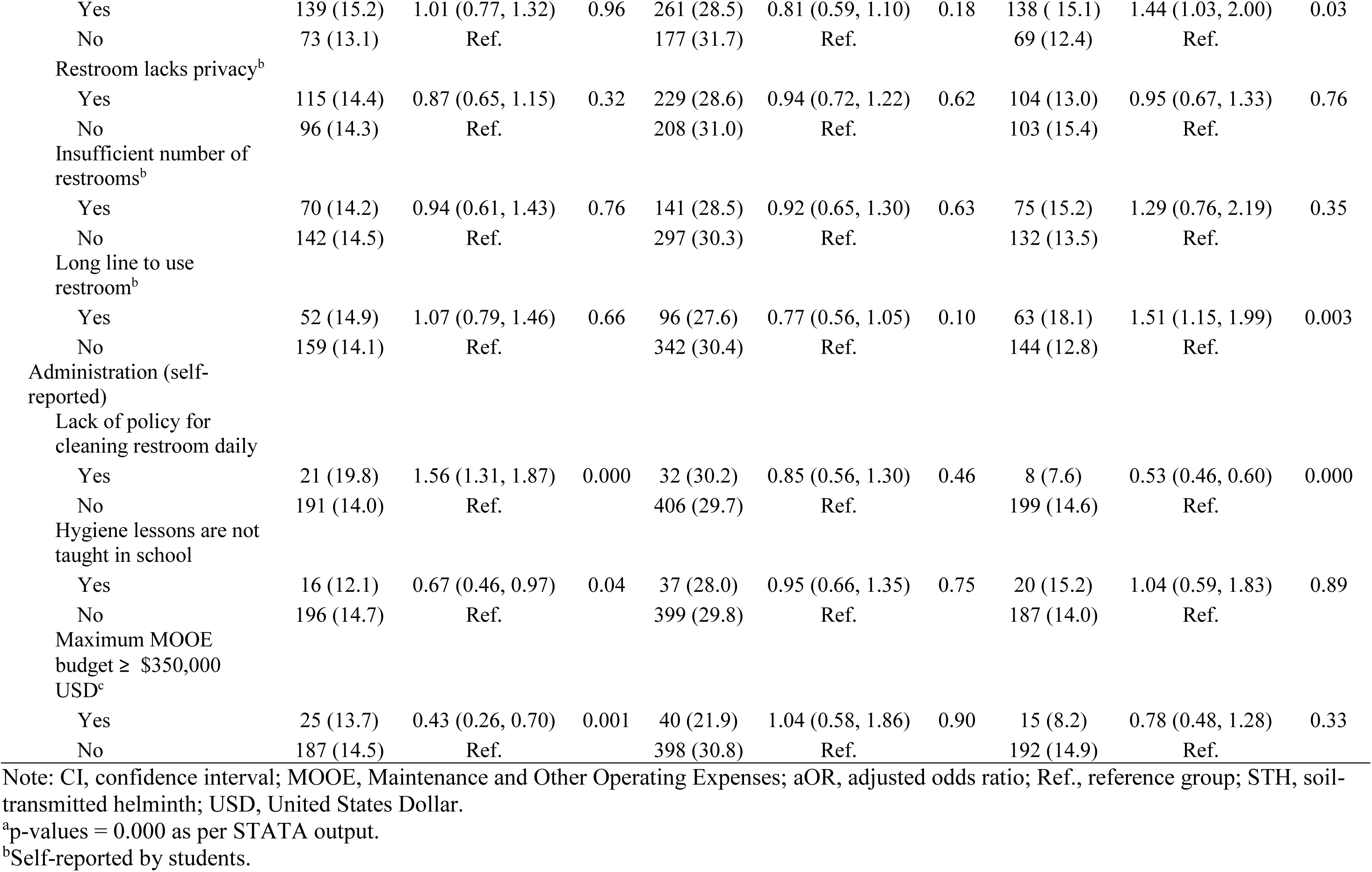
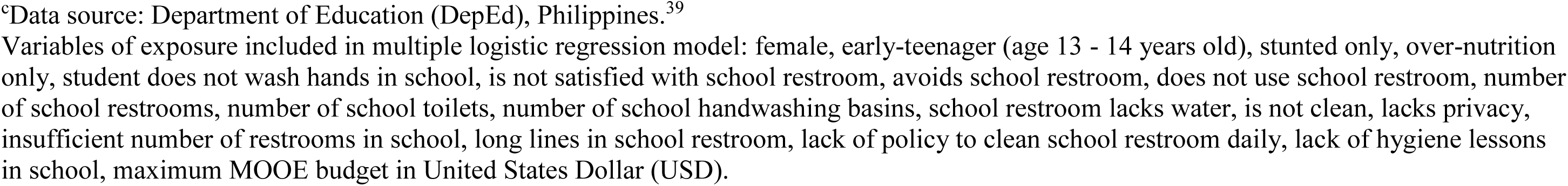
Multiple logistic regression models of self-reported diarrhea and STH infection among students (*N* = 1,296) from main sample and risk factors in schools (*N* = 14).

Students’ lack of satisfaction with school restrooms was significantly associated with increased odds of STH infection only, 1.40 (95% CI: 1.02, 1.90) (Table 5). The odds of STH infection decreased as the number of school WaSH facilities (e.g. restrooms, toilets, handwashing basins) increased. As school enrollment size increased, males’ odds of STH infection increased, while females’ odds decreased (Figure S1). We report results involving student-to-classroom ratios in Figure S2. Increased odds of dual infection were significantly associated with: restrooms’ insufficient water supply, 1.45 (95% CI: 1.16, 1.82); lack of cleanliness, 1.44 (95% CI: 1.03, 2.00); long lines to use the toilet, 1.51 (95% CI: 1.15, 2.00). An increasing number of school restrooms was significantly associated with decreased odds of dual infection (Table 5).

### Associations between Malnutrition and School WaSH

In multiple logistic regression models, significant risk factors of stunting only were: students’ diarrhea, 1.45 (95% CI: 1.04, 2.03); STH infection, 1.72 (95% CI: 1.17, 2.52); schools’ not providing hygiene lessons, 1.92 (95% CI: 1.14, 3.24) (Table 6). As school enrollment size increased, females’ odds of stunting tended to increase, while males’ odds tended to decrease (Figure S1). Schools’ having the maximum MOOE budget was significantly associated with decreased odds of stunting only, 0.25 (95% CI: 0.13, 0.50). Increased odds of undernutrition was significantly associated with: students’ STH infection, 1.50 (95% CI: 1.03, 2.19); schools’ insufficient number of restrooms for the number of users, 1.88 (95% CI: 1.04, 3.39); annual enrollment <= 2,000, 3.27 (95% CI: 1.84, 5.79) (Table. 6).

**Table 6.**
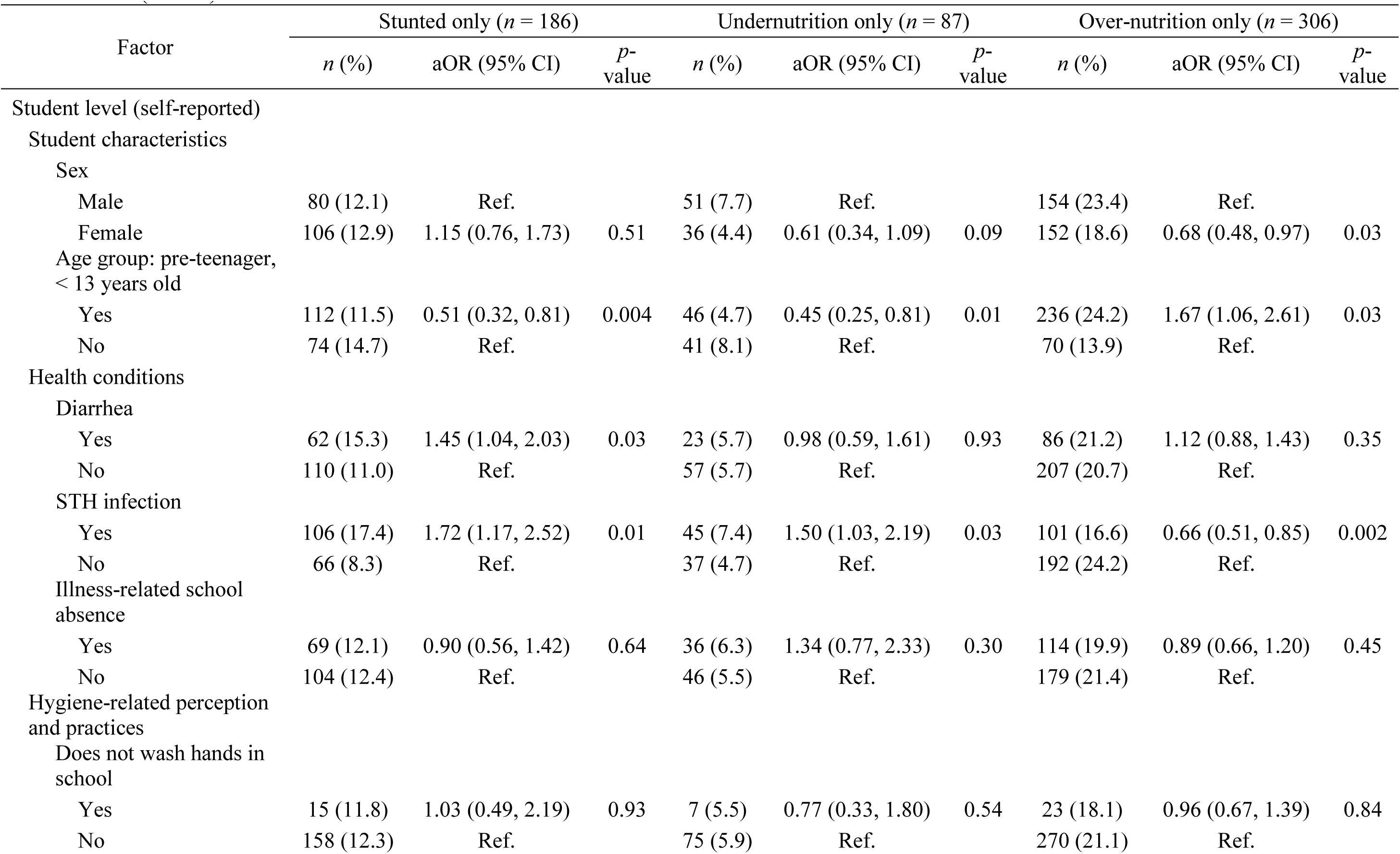
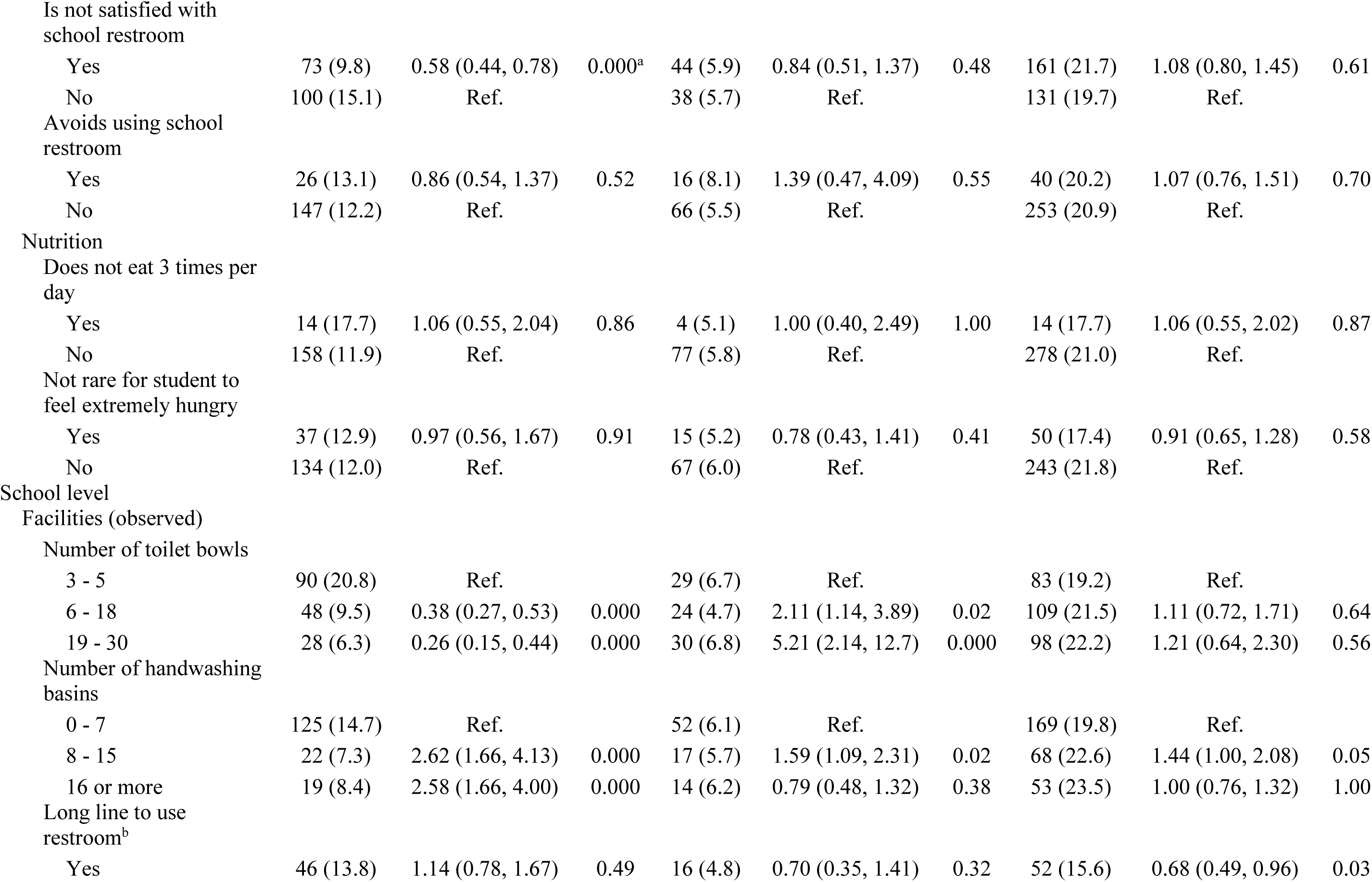
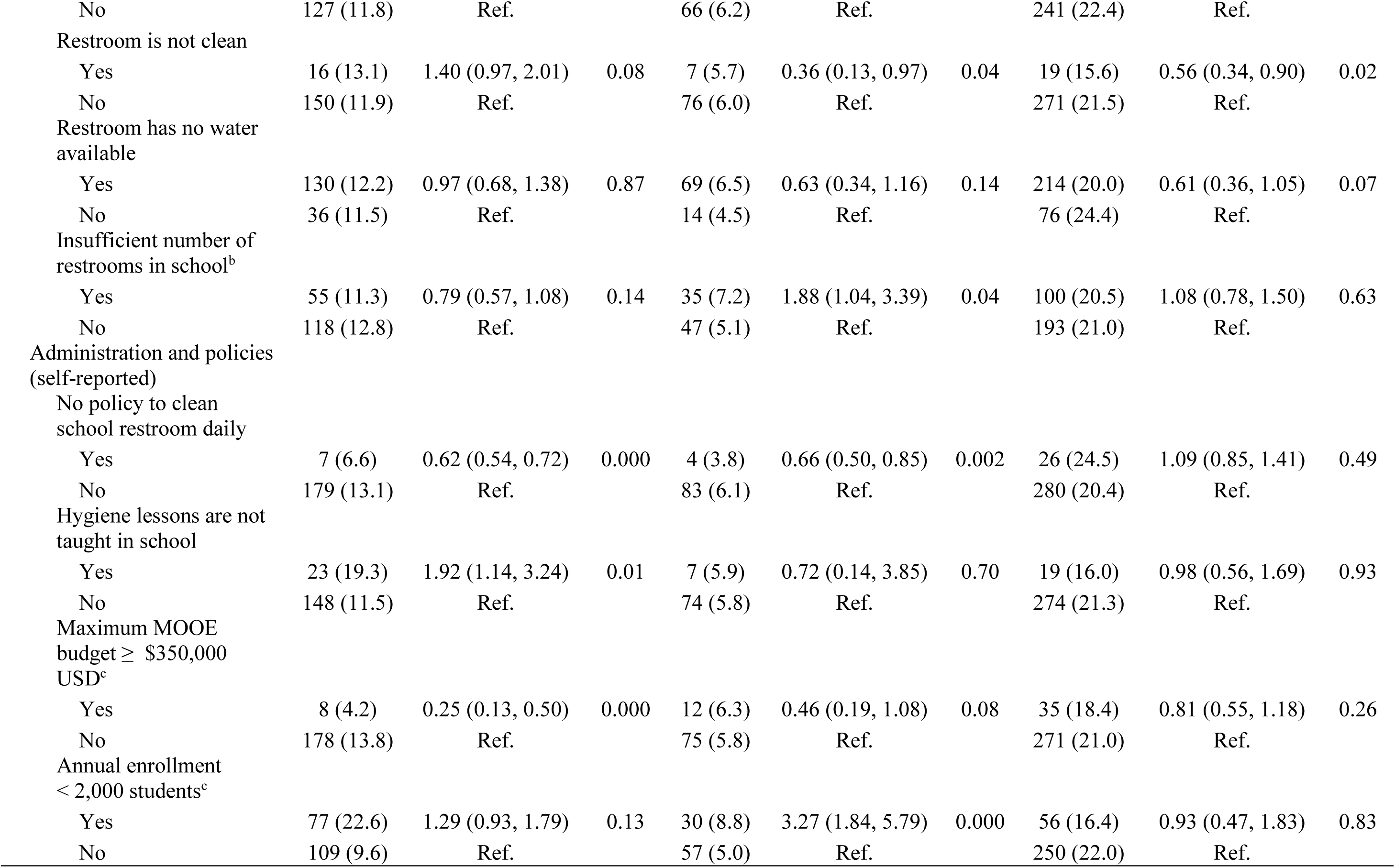
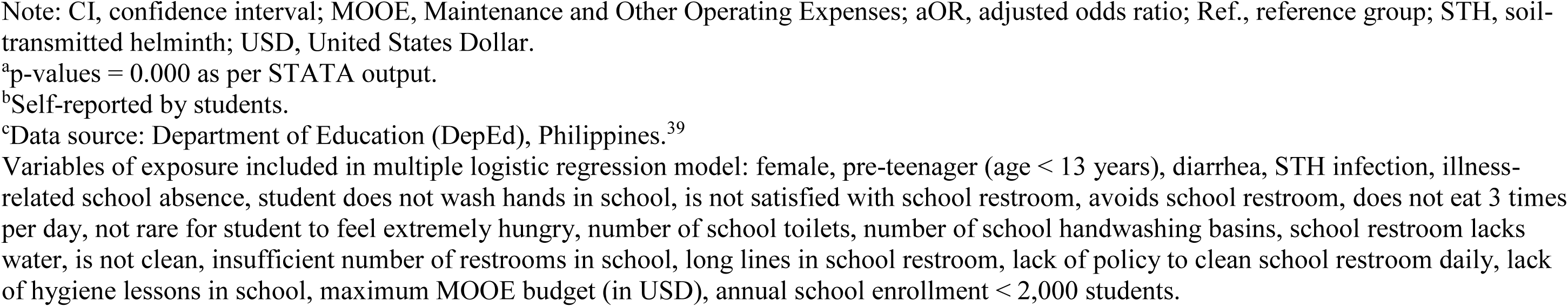
Multiple logistic regression model of observed malnutrition among students (*n* = 1,292) from main sample and risk factors in schools in Metro Manila (*n* = 14).

| Factor | Stunted only ($n = 186$) | | | Undernutrition only ($n = 87$) | | | Over-nutrition only ($n = 306$) | | |
| --- | --- | --- | --- | --- | --- | --- | --- | --- | --- |
| | $n$ (%) | aOR (95% CI) | $P$-value | $n$ (%) | aOR (95% CI) | $P$-value | $n$ (%) | aOR (95% CI) | $P$-value |
| Student level (self-reported) |  |  |  |  |  |  |  |  |  |
| Student characteristics |  |  |  |  |  |  |  |  |  |
| Sex |  |  |  |  |  |  |  |  |  |
| Male | 80 (12.1) | Ref. |  | 51 (7.7) | Ref. |  | 154 (23.4) | Ref. |  |
| Female | 106 (12.9) | 1.15 (0.76, 1.73) | 0.51 | 36 (4.4) | 0.61 (0.34, 1.09) | 0.09 | 152 (18.6) | 0.68 (0.48, 0.97) | 0.03 |
| Age group: pre-teenager, < 13 years old |  |  |  |  |  |  |  |  |  |
| Yes | 112 (11.5) | 0.51 (0.32, 0.81) | 0.004 | 46 (4.7) | 0.45 (0.25, 0.81) | 0.01 | 236 (24.2) | 1.67 (1.06, 2.61) | 0.03 |
| No | 74 (14.7) | Ref. |  | 41 (8.1) | Ref. |  | 70 (13.9) | Ref. |  |
| Health conditions |  |  |  |  |  |  |  |  |  |
| Diarrhea |  |  |  |  |  |  |  |  |  |
| Yes | 62 (15.3) | 1.45 (1.04, 2.03) | 0.03 | 23 (5.7) | 0.98 (0.59, 1.61) | 0.93 | 86 (21.2) | 1.12 (0.88, 1.43) | 0.35 |
| No | 110 (11.0) | Ref. |  | 57 (5.7) | Ref. |  | 207 (20.7) | Ref. |  |
| STH infection |  |  |  |  |  |  |  |  |  |
| Yes | 106 (17.4) | 1.72 (1.17, 2.52) | 0.01 | 45 (7.4) | 1.50 (1.03, 2.19) | 0.03 | 101 (16.6) | 0.66 (0.51, 0.85) | 0.002 |
| No | 66 (8.3) | Ref. |  | 37 (4.7) | Ref. |  | 192 (24.2) | Ref. |  |
| Illness-related school absence |  |  |  |  |  |  |  |  |  |
| Yes | 69 (12.1) | 0.90 (0.56, 1.42) | 0.64 | 36 (6.3) | 1.34 (0.77, 2.33) | 0.30 | 114 (19.9) | 0.89 (0.66, 1.20) | 0.45 |
| No | 104 (12.4) | Ref. |  | 46 (5.5) | Ref. |  | 179 (21.4) | Ref. |  |
| Hygiene-related perception and practices |  |  |  |  |  |  |  |  |  |
| Does not wash hands in school |  |  |  |  |  |  |  |  |  |
| Yes | 15 (11.8) | 1.03 (0.49, 2.19) | 0.93 | 7 (5.5) | 0.77 (0.33, 1.80) | 0.54 | 23 (18.1) | 0.96 (0.67, 1.39) | 0.84 |
| No | 158 (12.3) | Ref. |  | 75 (5.9) | Ref. |  | 270 (21.1) | Ref. |  |
| Is not satisfied with school restroom |  |  |  |  |  |  |  |  |  |
| Yes | 73 (9.8) | 0.58 (0.44, 0.78) | 0.000<sup>a</sup> | 44 (5.9) | 0.84 (0.51, 1.37) | 0.48 | 161 (21.7) | 1.08 (0.80, 1.45) | 0.61 |
| No | 100 (15.1) | Ref. |  | 38 (5.7) | Ref. |  | 131 (19.7) | Ref. |  |
| Avoids using school restroom |  |  |  |  |  |  |  |  |  |
| Yes | 26 (13.1) | 0.86 (0.54, 1.37) | 0.52 | 16 (8.1) | 1.39 (0.47, 4.09) | 0.55 | 40 (20.2) | 1.07 (0.76, 1.51) | 0.70 |
| No | 147 (12.2) | Ref. |  | 66 (5.5) | Ref. |  | 253 (20.9) | Ref. |  |
| Nutrition |  |  |  |  |  |  |  |  |  |
| Does not eat 3 times per day |  |  |  |  |  |  |  |  |  |
| Yes | 14 (17.7) | 1.06 (0.55, 2.04) | 0.86 | 4 (5.1) | 1.00 (0.40, 2.49) | 1.00 | 14 (17.7) | 1.06 (0.55, 2.02) | 0.87 |
| No | 158 (11.9) | Ref. |  | 77 (5.8) | Ref. |  | 278 (21.0) | Ref. |  |
| Not rare for student to feel extremely hungry |  |  |  |  |  |  |  |  |  |
| Yes | 37 (12.9) | 0.97 (0.56, 1.67) | 0.91 | 15 (5.2) | 0.78 (0.43, 1.41) | 0.41 | 50 (17.4) | 0.91 (0.65, 1.28) | 0.58 |
| No | 134 (12.0) | Ref. |  | 67 (6.0) | Ref. |  | 243 (21.8) | Ref. |  |
| School level |  |  |  |  |  |  |  |  |  |
| Facilities (observed) |  |  |  |  |  |  |  |  |  |
| Number of toilet bowls |  |  |  |  |  |  |  |  |  |
| 3 - 5 | 90 (20.8) | Ref. |  | 29 (6.7) | Ref. |  | 83 (19.2) | Ref. |  |
| 6 - 18 | 48 (9.5) | 0.38 (0.27, 0.53) | 0.000 | 24 (4.7) | 2.11 (1.14, 3.89) | 0.02 | 109 (21.5) | 1.11 (0.72, 1.71) | 0.64 |
| 19 - 30 | 28 (6.3) | 0.26 (0.15, 0.44) | 0.000 | 30 (6.8) | 5.21 (2.14, 12.7) | 0.000 | 98 (22.2) | 1.21 (0.64, 2.30) | 0.56 |
| Number of handwashing basins |  |  |  |  |  |  |  |  |  |
| 0 - 7 | 125 (14.7) | Ref. |  | 52 (6.1) | Ref. |  | 169 (19.8) | Ref. |  |
| 8 - 15 | 22 (7.3) | 2.62 (1.66, 4.13) | 0.000 | 17 (5.7) | 1.59 (1.09, 2.31) | 0.02 | 68 (22.6) | 1.44 (1.00, 2.08) | 0.05 |
| 16 or more | 19 (8.4) | 2.58 (1.66, 4.00) | 0.000 | 14 (6.2) | 0.79 (0.48, 1.32) | 0.38 | 53 (23.5) | 1.00 (0.76, 1.32) | 1.00 |
| Long line to use restroom<sup>b</sup> |  |  |  |  |  |  |  |  |  |
| Yes | 46 (13.8) | 1.14 (0.78, 1.67) | 0.49 | 16 (4.8) | 0.70 (0.35, 1.41) | 0.32 | 52 (15.6) | 0.68 (0.49, 0.96) | 0.03 |
| No | 127 (11.8) | Ref. |  | 66 (6.2) | Ref. |  | 241 (22.4) | Ref. |  |
| Restroom is not clean |  |  |  |  |  |  |  |  |  |
| Yes | 16 (13.1) | 1.40 (0.97, 2.01) | 0.08 | 7 (5.7) | 0.36 (0.13, 0.97) | 0.04 | 19 (15.6) | 0.56 (0.34, 0.90) | 0.02 |
| No | 150 (11.9) | Ref. |  | 76 (6.0) | Ref. |  | 271 (21.5) | Ref. |  |
| Restroom has no water available |  |  |  |  |  |  |  |  |  |
| Yes | 130 (12.2) | 0.97 (0.68, 1.38) | 0.87 | 69 (6.5) | 0.63 (0.34, 1.16) | 0.14 | 214 (20.0) | 0.61 (0.36, 1.05) | 0.07 |
| No | 36 (11.5) | Ref. |  | 14 (4.5) | Ref. |  | 76 (24.4) | Ref. |  |
| Insufficient number of restrooms in school<sup>b</sup> |  |  |  |  |  |  |  |  |  |
| Yes | 55 (11.3) | 0.79 (0.57, 1.08) | 0.14 | 35 (7.2) | 1.88 (1.04, 3.39) | 0.04 | 100 (20.5) | 1.08 (0.78, 1.50) | 0.63 |
| No | 118 (12.8) | Ref. |  | 47 (5.1) | Ref. |  | 193 (21.0) | Ref. |  |
| Administration and policies (self-reported) |  |  |  |  |  |  |  |  |  |
| No policy to clean school restroom daily |  |  |  |  |  |  |  |  |  |
| Yes | 7 (6.6) | 0.62 (0.54, 0.72) | 0.000 | 4 (3.8) | 0.66 (0.50, 0.85) | 0.002 | 26 (24.5) | 1.09 (0.85, 1.41) | 0.49 |
| No | 179 (13.1) | Ref. |  | 83 (6.1) | Ref. |  | 280 (20.4) | Ref. |  |
| Hygiene lessons are not taught in school |  |  |  |  |  |  |  |  |  |
| Yes | 23 (19.3) | 1.92 (1.14, 3.24) | 0.01 | 7 (5.9) | 0.72 (0.14, 3.85) | 0.70 | 19 (16.0) | 0.98 (0.56, 1.69) | 0.93 |
| No | 148 (11.5) | Ref. |  | 74 (5.8) | Ref. |  | 274 (21.3) | Ref. |  |
| Maximum MOOE budget ≥ \$350,000 USD<sup>c</sup> | | | | | | | | | |
| Yes | 8 (4.2) | 0.25 (0.13, 0.50) | 0.000 | 12 (6.3) | 0.46 (0.19, 1.08) | 0.08 | 35 (18.4) | 0.81 (0.55, 1.18) | 0.26 |
| No | 178 (13.8) | Ref. |  | 75 (5.8) | Ref. |  | 271 (21.0) | Ref. |  |
| Annual enrollment < 2,000 students<sup>c</sup> |  |  |  |  |  |  |  |  |  |
| Yes | 77 (22.6) | 1.29 (0.93, 1.79) | 0.13 | 30 (8.8) | 3.27 (1.84, 5.79) | 0.000 | 56 (16.4) | 0.93 (0.47, 1.83) | 0.83 |
| No | 109 (9.6) | Ref. |  | 57 (5.0) | Ref. |  | 250 (22.0) | Ref. |  |

Note: CI, confidence interval; MOOE, Maintenance and Other Operating Expenses; aOR, adjusted odds ratio; Ref., reference group; STH, soil-transmitted helminth; USD, United States Dollar.
<sup>a</sup>p-values = 0.000 as per STATA output.
<sup>b</sup>Self-reported by students.
<sup>c</sup>Data source: Department of Education (DepEd), Philippines.<sup>39</sup>
Variables of exposure included in multiple logistic regression model: female, pre-teenager (age < 13 years), diarrhea, STH infection, illness-related school absence, student does not wash hands in school, is not satisfied with school restroom, avoids school restroom, does not eat 3 times per day, not rare for student to feel extremely hungry, number of school toilets, number of school handwashing basins, school restroom lacks water, is not clean, insufficient number of restrooms in school, long lines in school restroom, lack of policy to clean school restroom daily, lack of hygiene lessons in school, maximum MOOE budget (in USD), annual school enrollment < 2,000 students.

### Associations with Subsample of Students’ Households

#### Impact of Home Level Factors on Diarrhea, STH infection, and Malnutrition

In multiple logistic regression models, home level factors non-significantly associated with increased odds of diarrhea only were: no restroom inside the home, 1.82 (95% CI: 0.36, 9.28); no water supply in restroom, 3.63 (95% CI: 0.30, 43.9); no handwashing basin in restroom, 1.28 (95% CI: 0.20, 8.39) (Table 7). Of note, the above results had 95% CIs that included the value of ‘1’; thus, we cannot be completely sure if the odds increase or decrease.^68^ However, because of asymmetry in the 95% CIs, leaning in the positive direction, we can estimate that the odds increase rather than decrease. Increased odds of STH infection only was significantly associated with having no water supply in the restroom, 32.2 (95% CI: 2.52, 411.0).

**Table 7.**
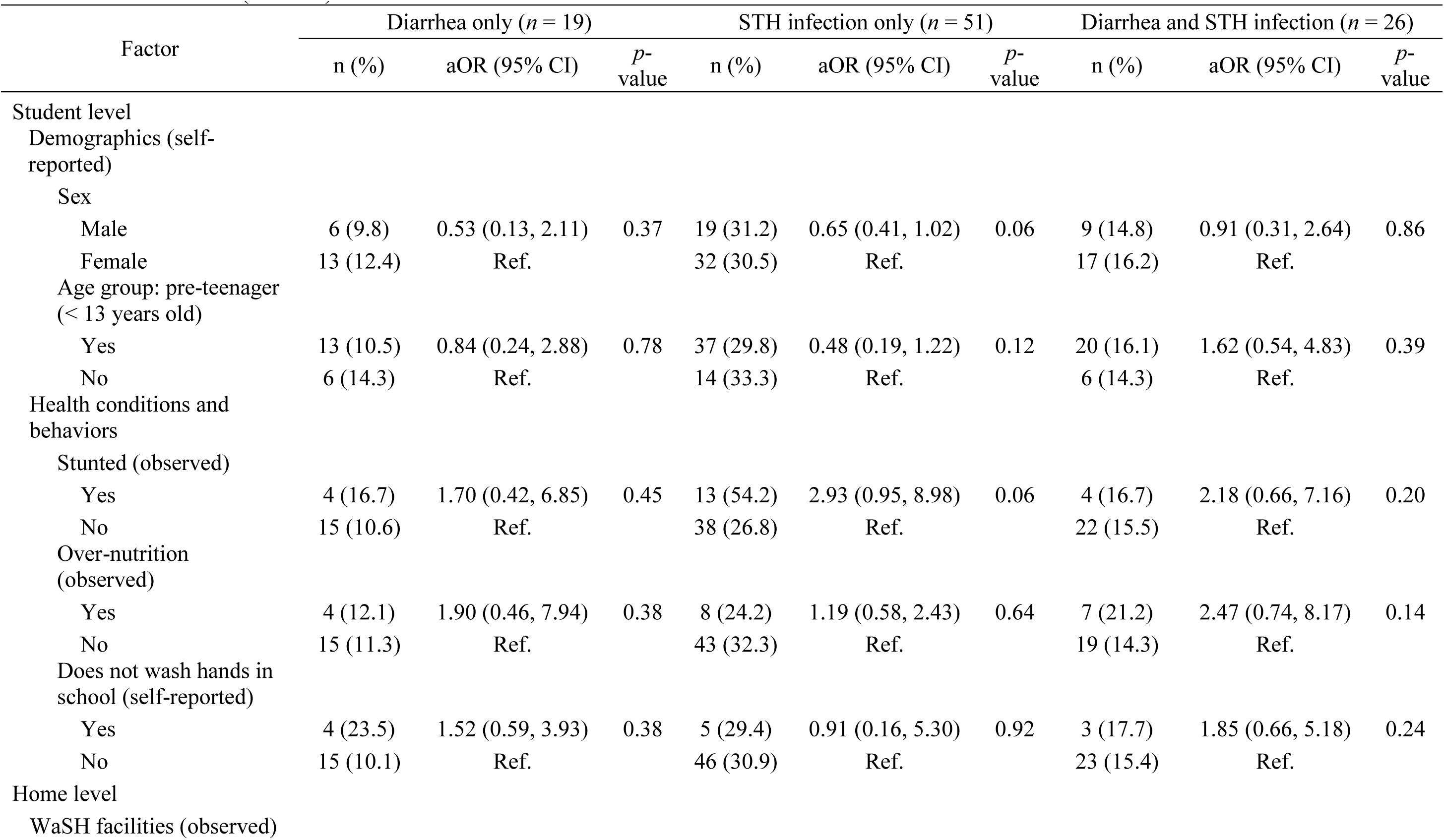
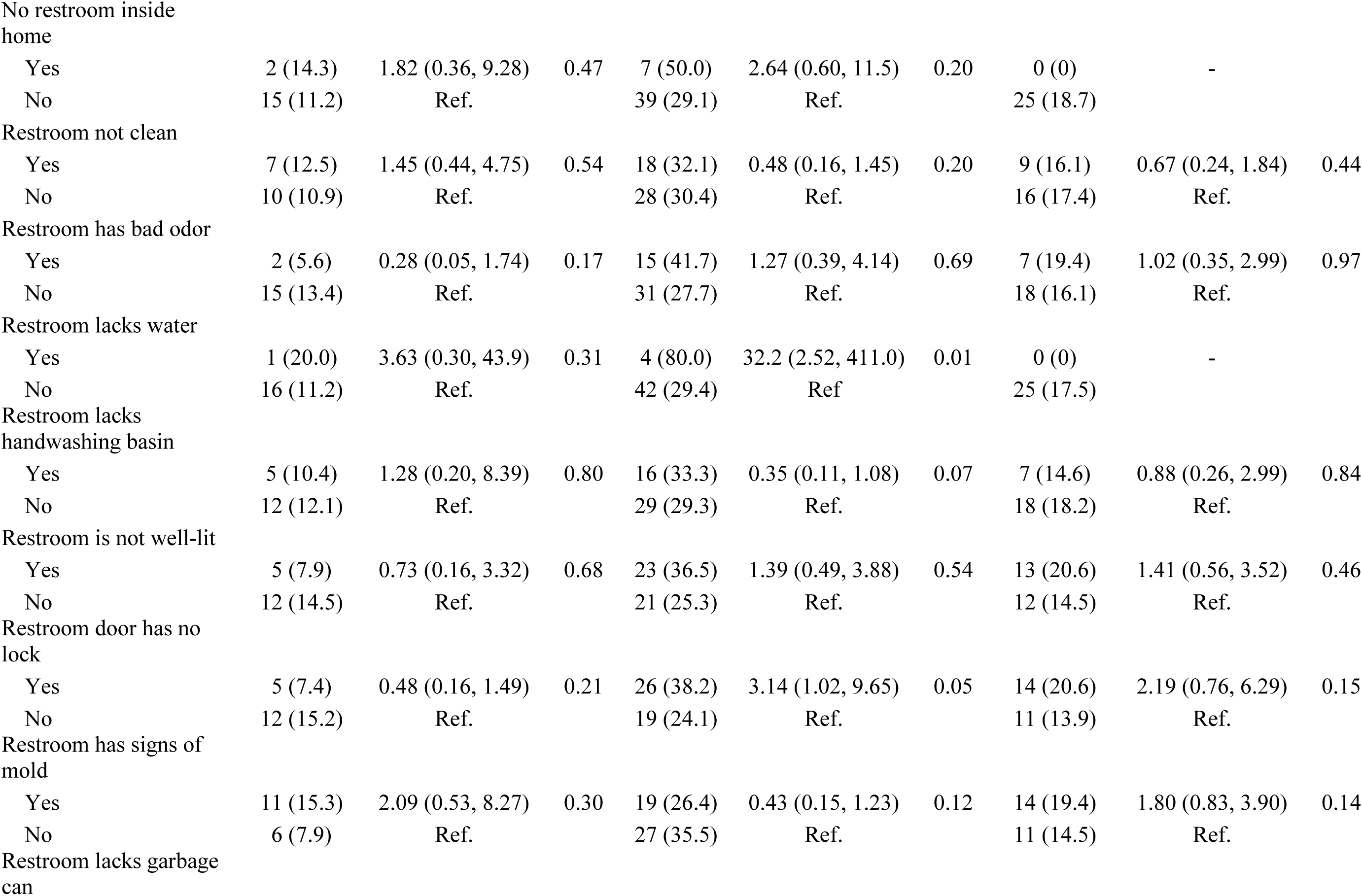
Multiple logistic regression models of self-reported diarrhea and STH infection among students from subsample and risk factors in homes in Metro Manila (*n* = 212).

Home restrooms’ having signs of mold, 3.76 (95% CI: 1.78, 7.94), and having no garbage can, 3.50 (95% CI: 1.77, 6.91), were significantly associated with increased odds of stunting only (Table 8). Home-level factors non-significantly associated with increased odds of undernutrition were: families’ experiences with asking/begging for food, 4.75 (95% CI: 0.23, 98.1); eating precooked food more often than freshly cooked food, 3.69 (95% CI: 0.94, 14.6); restroom has signs of mold, 1.05 (95% CI: 0.38, 2.93). Again, the above results had 95% CIs that included the value of ‘1’. Thus, while we cannot be completely sure if the odds increase or decrease, the asymmetry in the 95% CIs, leaning in the positive direction, leads us to estimate that the odds increase rather than decrease. Increased odds of STH infection only was significantly associated with having no water supply in the restroom, 32.2 (95% CI: 2.52, 411.0).

**Table 8.**
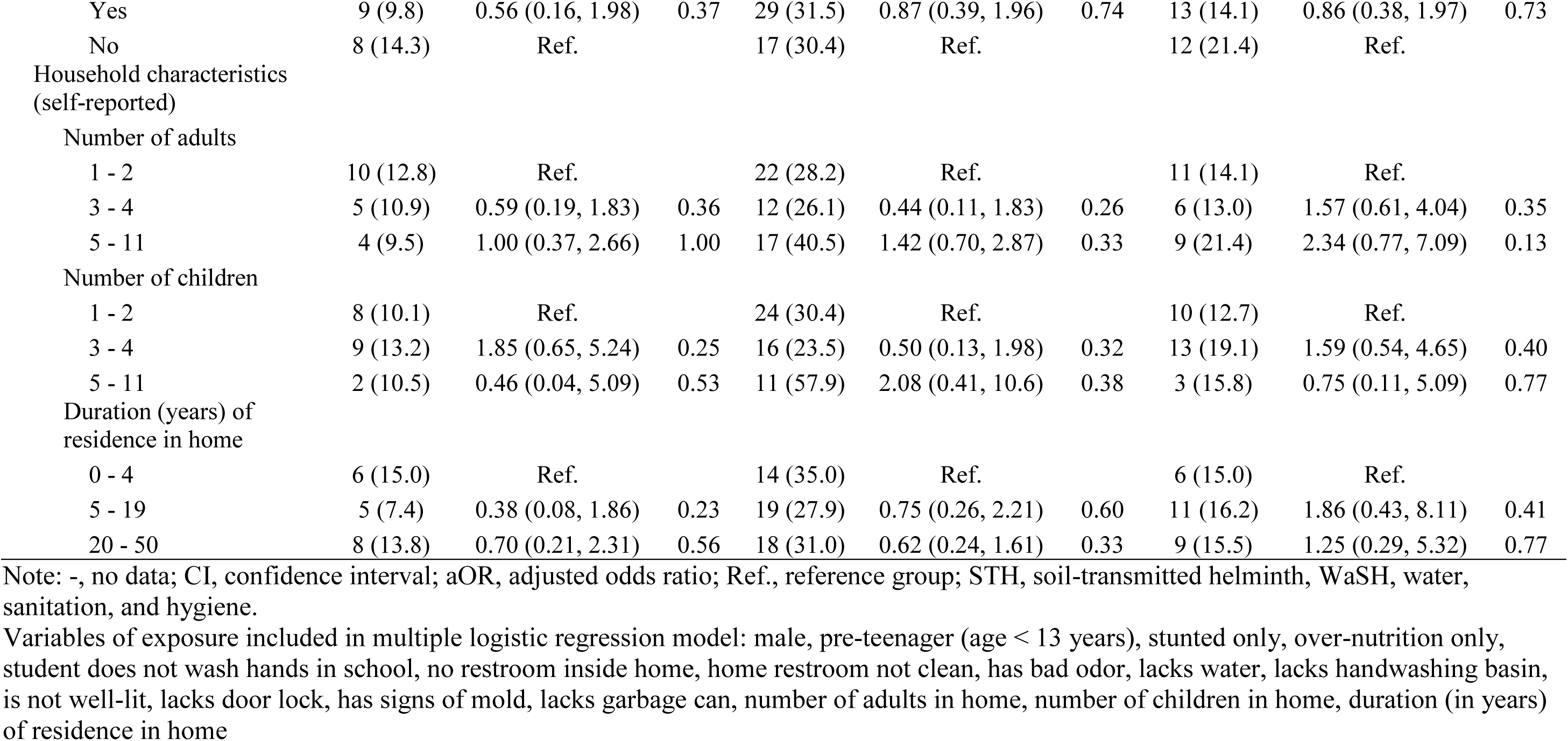
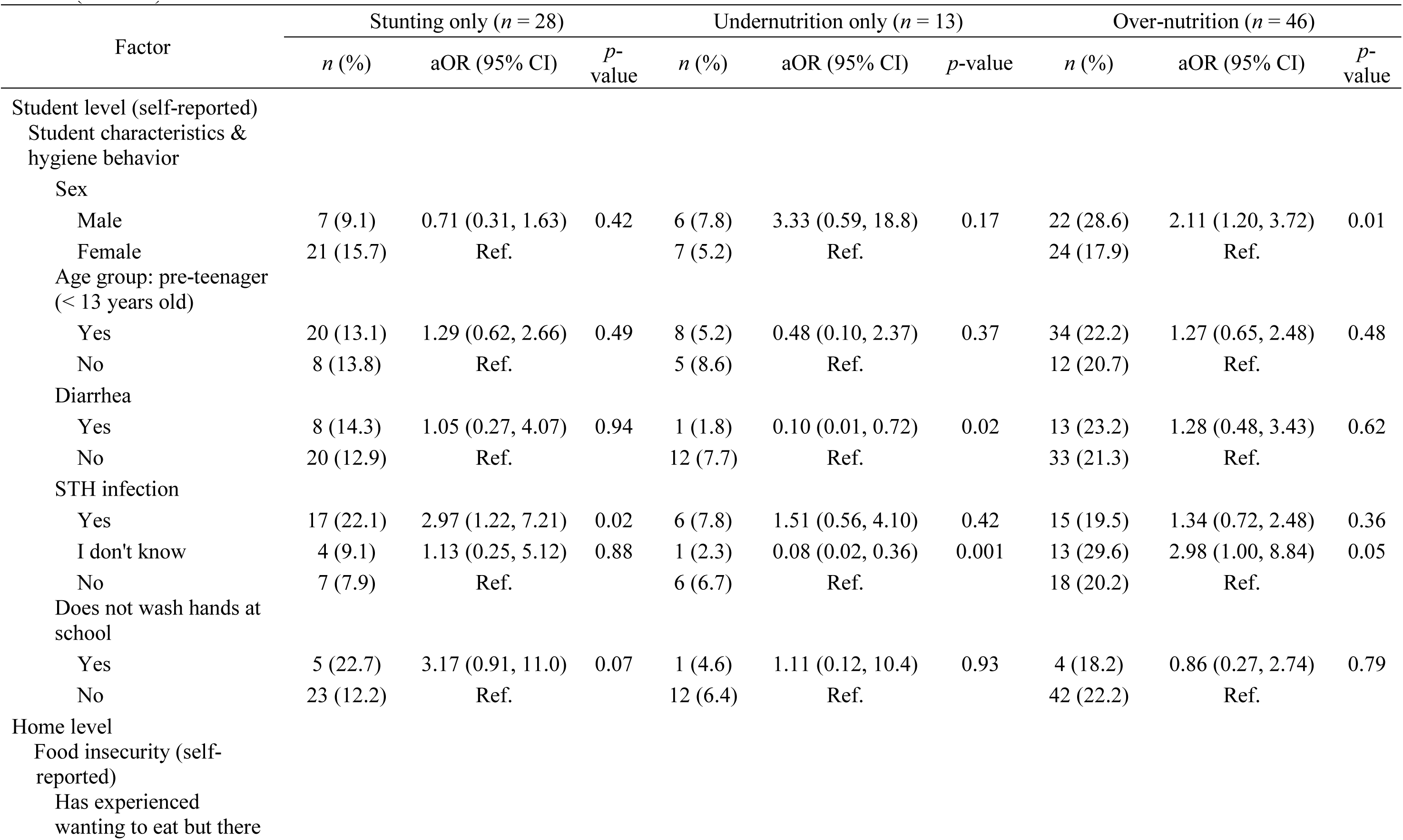
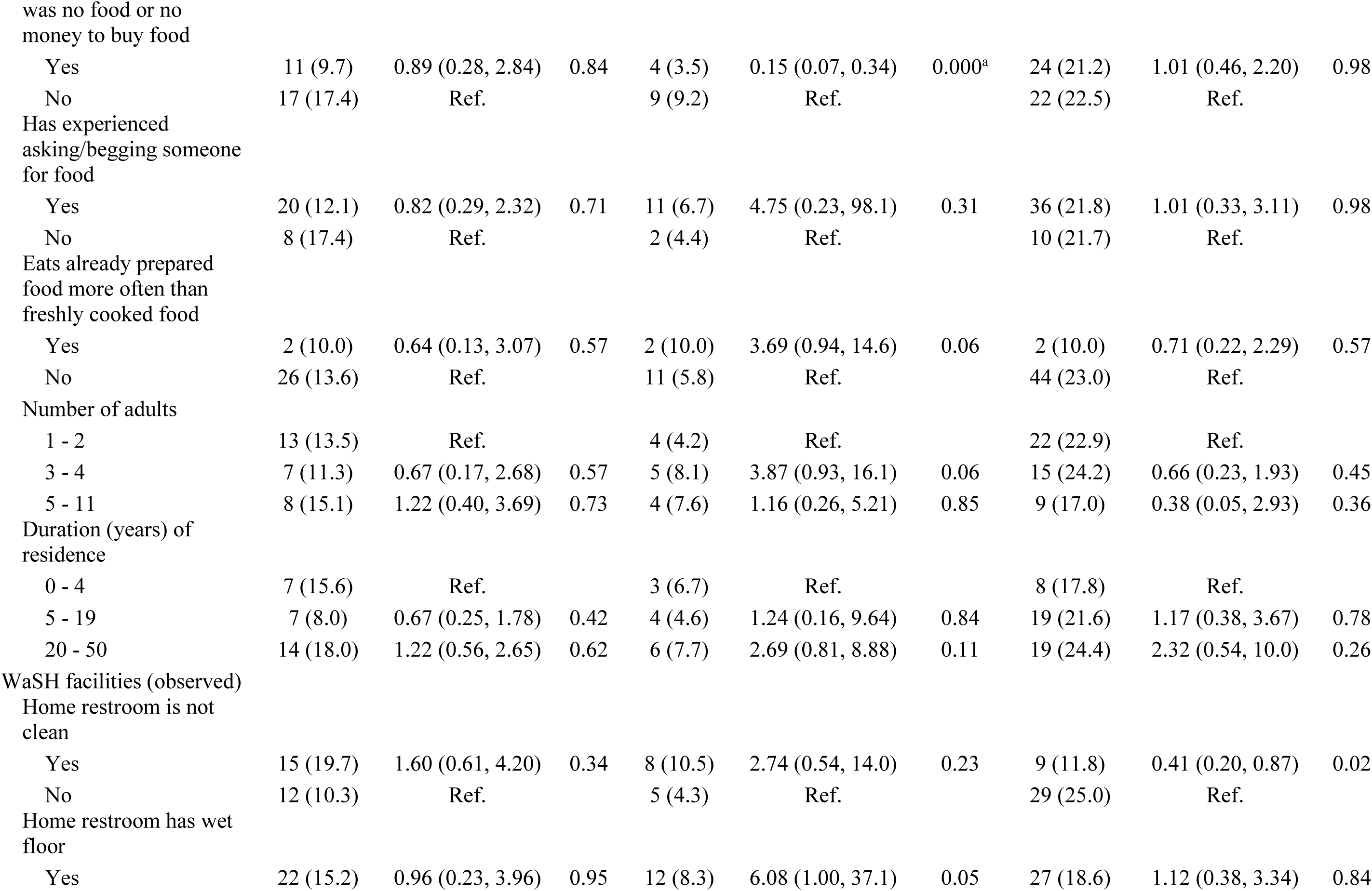
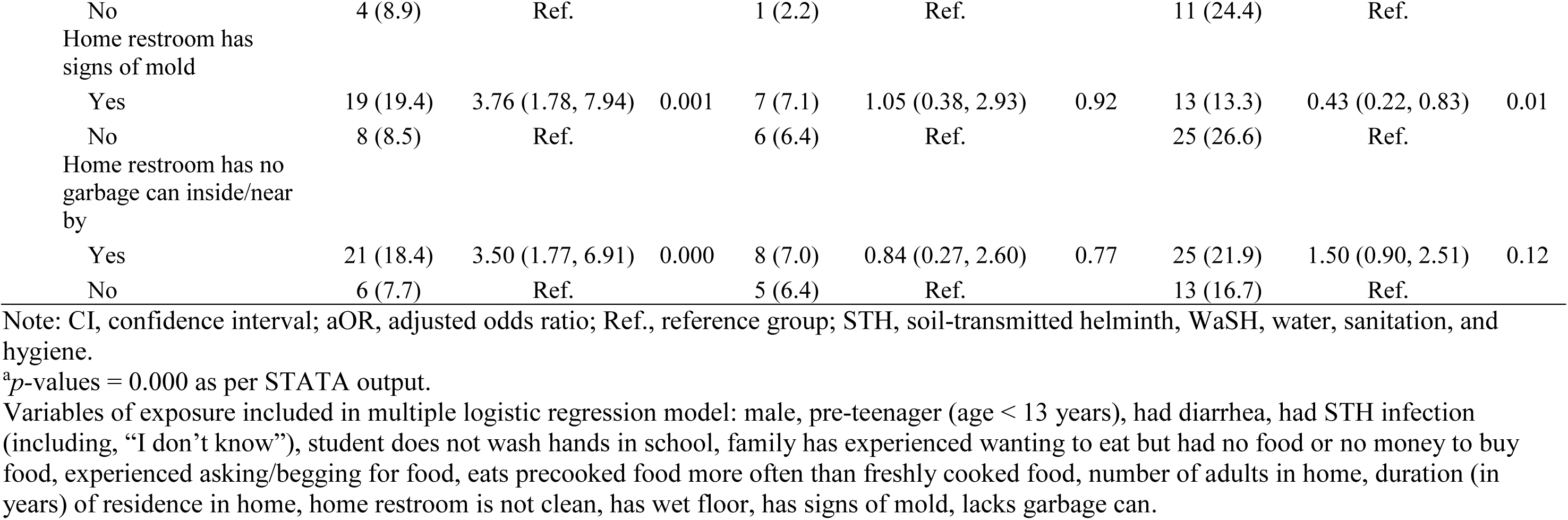
Multiple logistic regression models of observed malnutrition among students from subsample and risk factors in homes in Metro Manila (*n* = 211).

## Discussion

### Key Findings and Interpretation

Diarrhea and STH infection were highly prevalent in our sample: about 3 out of 4 students (72.5%, 1,068) had at least one of the infections during the specified time period. Disease prevalence rates from our study were higher compared to those from previous studies of schoolchildren with diarrhea or STH infection in low-income settings.^40, 69, 70^ Our findings indicate that schools’ insufficient number of toilets and inadequate maintenance of restrooms, coupled with students’ poor handwashing, are increasing the risk of diarrhea and STH infection. In schools with 3-5 toilets, students reported significantly greater odds of diarrhea and non-significantly greater odds of STH infection. In schools with 6-18 toilets, students were also more likely to report both of these outcomes but with a smaller aOR, suggesting that disease risk decreases as the number of toilets increases. Furthermore, poor handwashing was associated with increased risk of diarrhea only, demonstrating how crucial it is to provide adequate handwashing facilities and lessons in schools to reduce diarrhea prevalence. Over half of students were not satisfied with school restrooms, with ∼2/3 of students reporting that restrooms were not clean and > 1/2 of students reporting that restrooms did not provide enough privacy. These findings were concordant with observations from research assistants. Yet in spite of school restrooms’ “bad” condition, almost all students still used the restrooms rather than practice open defecation. Furthermore, students who avoided using the restroom had a significantly greater risk of diarrhea. Thus, more investments are needed to improve and maintain school restrooms in order to promote their use by children.

The rate of self-reported handwashing was very high (91.3%), similar to previous studies in Indonesia^71^ and Kenya,^70^ although these studies occurred in homes rather than schools. Our study participants’ self-reported rate of handwashing with soap was 53%, which is lower than self-reported rates from similar studies in Indonesia^72^ and Vietnam.^73^ Our findings might be reflecting differences in collecting data via self-report, a method that has been associated with over-reporting, by as much as 50%-60%, compared to observed handwashing rates.^74^ We found an association between poor handwashing and increased risk of diarrhea only, STH infection only, both diarrhea and STH infection, and stunting. These findings contribute to the growing body of evidence that the promotion of handwashing in schools has many benefits, e.g. decreasing the prevalence of: diarrhea,^75^ diarrhea-related school absences,^76^ moderate to severe STH infection^77^ STH reinfection^78^ and stunting.^79^ The low rate of handwashing with soap found in our study may be attributed not only to students’ behavior or lack of knowledge, but also to the lack of soap provided by schools. Such a finding is of concern, considering that community interventions using handwashing with soap, assessed via observation, have reduced the risk of diarrheal diseases by up to 47%.^80^

An unexpected, non-significant association was found between schools’ not providing hygiene lessons and decreased odds of diarrhea only and STH infection only. One reason could be the presence of unmeasured confounders. Another reason is the complicated relationship between exposures and outcomes. Multiple sectors overlap in WaSH management, making it more difficult to pinpoint or address risk factors of diarrhea and STH infection. This may also explain the unexpected association we found between schools’ not having a policy for cleaning the restroom daily and decreased odds of STH infection only and decreased odds of both diarrhea and STH infection. However, schools’ lack of policy for cleaning the restroom daily was significantly associated with increased odds of diarrhea only. The school principals we interviewed almost unanimously reported that WaSH management policies were in place, yet the “bad” conditions of school restrooms reported by students and observed by researcher assistants begged the question of whether or not the policies were being effectively enforced. We found an unexpected, non-significant association between schools’ having the maximum MOOE budget and increased odds of STH infection. However, this association disappeared when we estimated the odds of STH infection in primary and secondary schools separately. Instead, we found a significant association between schools’ having the maximum MOOE budget and decreased odds of STH infection. These findings may point to possible gaps in resource utilization, incident reporting, auditing, or communication, or factors outside of schools.

We assessed water security in two ways: 1) by asking children, with one item in the questionnaire, if water was available in the school restrooms; and 2) by inspecting school restrooms for available water. About 22% of children reported that no water was available in school restrooms. About 33% of restrooms were observed to have had no water available.

The lack of water was significantly associated with increased odds of dual infection (both diarrhea and STH infection), but it was also unexpectedly associated with decreased odds of diarrhea only (Table 5). One possible explanation for this: If water was heavily contaminated with *E.coli* and coliform, then having no water in the school restrooms could actually be a protective factor, as children would have decreased exposure to water that contained diarrhea-causing microorganisms. As a result, children’s risk for diarrhea would be decreased and we would see decreased diarrhea prevalence. Another explanation could be that other routes of disease transmission are more important than quantity or quality of water for this population, as previously described by Weaver and others,^81^ citing Moe and others,^82^ who reported that Filipino children who drank moderately contaminated water had similar diarrheal disease rates as those who drank uncontaminated water. Nevertheless, the availability of water in school restrooms remains an important factor that should not be ignored. We found a significant association between school restrooms’ lack of water and increased odds of STH infection and stunting when we estimated odds of disease in primary and secondary schools separately.

It is unclear how students’ attitudes and behaviors impact diarrhea and STH infection. Students’ lack of satisfaction with restrooms was significantly associated with increased odds of STH infection only but with decreased odds of dual infection (both diarrhea and STH infection). Students’ avoidance of restrooms was significantly associated with increased odds of diarrhea but with decreased odds of STH infection. These mixed results suggest that unmeasured factors likely exist and are confounding the relationship between exposures and outcomes.

Younger teenagers (13-14 years old) were more likely to report all 3 disease outcomes compared to their peers from other age groups. Our finding of higher diarrhea and STH infection prevalence rates in younger compared to older children was consistent with some previous studies,^83, 84^ but in contrast with other studies.^85, 86^ Some reasons may be younger children’s lower levels of immunity, increased exposures to risk factors from the fecal-oral route, or limited capacity to maintain hygiene practices over time.

Over-crowding of students on school campuses may be another factor increasing the risk of disease, and the severity of health consequences may be sex-specific. As schools’ annual enrollment size increased, we found the odds of diarrhea only increased in females but decreased in males. Females’ odds of diarrhea only increased as the student-to-classroom ratio increased. Females’ odds of STH infection only decreased as the student-to-classroom ratio increased from the 4^th^ to the 5^th^ highest student-to-classroom ratio categories, regardless of the number of school-day shifts (single or double). In contrast, males seemed to be more at risk of STH infection as over-crowding increased. Our findings were in line with those from previous studies that identified over-crowding as a risk factor for the spread of infectious diseases.^87, 88^ This emphasizes the need for sex-specific or gender-sensitive interventions, especially in over-crowded schools in the Global South.

We used multiple logistic regression models to assess risk factors of select health outcomes. We obtained results that had 95% CIs that included the value of “1”, which indicated that 1) the aOR was not statistically significant and 2) the aOR includes the likelihood for either an increase or reduction in the estimates of effect/outcome. To assess the likelihood that the direction of effect could be an increase or reduction, we looked at the 95% CI to see if the interval was symmetric. All results that contain the value of “1” in the 95% CI must be interpreted with caution as there is a likelihood of either an increase or reduction in the estimate/outcome.

### Limitations

Our study, like all observational studies, is limited by confounding. Because we did not use randomization, it is possible that our findings were affected by known (but uncontrolled for) confounders and unknown confounders. Possible consequences of confounding are: identifying (apparent) associations where none actually exist, ignoring associations that actually exist, and/or over-/underestimating the magnitude of observed associations. It was not possible for us to identify and control for all confounding variables in our study, e.g. environmental pollution surrounding school campuses, dietary patterns of children. Another example is SES, which is typically associated with enrollment in public school in the Philippines, infectious disease and malnutrition prevalence, and food insecurity. We did not assess parents’ educational background, employment, or household income; thus, our ability to control for confounding related to such variables was limited. In a forthcoming paper we will report findings from our longitudinal study, also part of the project, “WaSH in Manila Schools”, which took place in the same public schools and with a similar sample of children, where we did adjust for such covariates.

Our ability to interpret associations may be limited because we used a cross-sectional study design, i.e. measuring outcomes only once, rather than measuring outcomes >= 2 times, comparing change over time. We measured outcomes in one group only, rather than measuring and comparing outcomes between a control group and an experimental group. Our observational study enabled us to describe associations but not causality. Our study findings may be context-specific in terms of disease prevalence and environmental risk factors, which limit the generalizability of observed associations to similar settings and populations, e.g. urban poor schoolchildren living in LMICs located in the tropics.

Prior to conducting school surveys, selection bias was likely introduced to our study, as we excluded from our sampling frame schools with a smaller enrollment size and we accepted one school from a third city that asked us directly to participate in the study. Allowing school principals/representatives to select which class section(s) would be surveyed may have limited the representativeness of study findings. This is because class sections were usually organized according to students’ academic performance. Often, top-performing students were grouped together in the first class sections (e.g. 1-2), while poor-performing students were grouped together in the last class sections (e.g. 9-10). We included in our analysis data from a school where we conducted pilot testing of our survey instruments. At this school, admission is based on admission test scores, and consideration is given to children whose parent works at the University of the Philippines. Some children attending this school may have a sociodemographic background that differs from that of children attending the other public schools we surveyed.

Risks of bias likely increased during the implementation of our study. First, recall bias likely increased when we used self-report to measure multiple outcomes. We used children’s self-reported diarrhea and STH infection data but did not corroborate with medical records. The accuracy of self-reported data depended on children’s ability to understand the case definitions we provided for diarrhea and STH infection, their ability to remember whether or not they had symptoms indicative of these diseases, their willingness to report these diseases, and their ability to correctly operate electronic tablets to complete our questionnaire. Children’s willingness to report diseases could have been influenced, on the one hand, by embarrassment or fear of repercussions, and on the other hand, by the desire for special attention or the wish to receive compensation. Possible reasons for children’s under-reporting of disease outcomes included: feeling too embarrassed, being scared that they or their parents would be stigmatized or punished, and not recognizing subtle symptoms of diarrhea or STH infection. Also, older children and adolescents may have forgotten about their STH infection if it occurred many years ago. Possible reasons for children’s over-reporting of disease outcomes included: desiring to receive extra attention, hoping to receive special treatment (i.e. exemption from attending school due to a medical condition), and hoping to receive compensation, free medication, or other resources for themselves, their family, or their school.

Children may have been motivated by different reasons to report their hygiene behaviors and perceptions about schools’ WaSH facilities. For example, they may have felt motivated to give socially “desirable” answers to avoid embarrassment, e.g. reporting that they regularly washed their hands after using the toilet. Other children may have under-reported the poor conditions of schools’ WaSH facilities to not bring “shame” or “dishonor” to their school or to school personnel. In contrast, some children could have over-reported their avoidance of the school restroom and dissatisfaction with schools’ WaSH facilities because they hoped money or other resources would be donated to the school for WaSH facilities improvements. Another possible reason for over-reporting was children’s desire to retaliate against school personnel.

Another possible source of bias was children’s self-reported avoidance of school restrooms and self-reported perceptions about school WaSH facilities. The rationale for using self-report were: reliability, convenience, feasibility, and the ability to quickly, affordably, and accurately assess a large study sample. Our use of self-report is supported by previous studies.^89–92^ It was important for us to understand children’s hygiene behaviors and practices. Indeed, we could have used alternative measurements, e.g. observing children physically avoid the restroom or interviewing teachers or other school personnel about their knowledge or perceptions about children’s avoidance of school restrooms. However, the ideal way to measure this outcome was to ask children directly. To help ensure that children gave honest answers, without feeling pressured to give “socially desirable” answers, we administered a self-administered questionnaire anonymously and maintained confidentiality. Children answered questionnaires privately, away from the gaze of classmates and school personnel. Researchers were trained to position themselves at some distance away from children and to not look directly at children’s answers. Researchers did not intervene during the questionnaire, unless children asked for assistance. Children were also told that their answers would not be shared with parents or school personnel, nor would their answers affect their school grades. It was important for us to assess school WaSH from the perspective of users (i.e. schoolchildren). We offered no compensation to school personnel or children in exchange for participating in our study. We promised no material reward or supplies for the school or children in exchange for participating in our study. The reason for the above measures was to make children feel comfortable about giving their honest answers without undue pressure. Furthermore, we compared data on schools’ WaSH facilities with two other sources of information: interviews with school principals and visual inspections of school restrooms conducted by our research team.

Second, outcome misclassification bias likely increased when we used some outcome measures that had limited reliability and/or validity. We did not use objective diagnostic data to measure the primary outcomes of diarrhea and STH infection prevalence. Instead, we used children’s self-report, which relies heavily on recall and is vulnerable to bias. A possible consequence is that some of the children’s self-reported disease outcomes could be inaccurate as they were not corroborated with clinical/diagnostic assessment.

Self-reported health outcomes are commonly used in epidemiologic studies when clinically confirming infections is not feasible. Self-reported health outcome data depended on children’s ability to recognize a disease based on symptoms, without a clinical diagnosis or verification by diagnostic testing. Self-report also depends on children’s ability to accurately recall possibly very subtle, or at least not very bothersome, clinical signs that appeared recently or not so recently. Using longer recall periods, e.g. from 6 months to one year, may be disadvantageous because children could forget to report important information. Self-report could also be influenced by children’s perceptions about what is “desirable” and “undesirable” to report because of social/cultural norms, feelings of shame, and fears about being punished or singled out, and, conversely, the desire to receive attention, special treatment, and resources for one’s family or school. Our study involved older school-age children and adolescents who, because of their developmental status, were more likely to be motivated by the desire to fit in with their peers and avoid embarrassment. Thus, we expect that they were less likely to over-report “problematic” conditions or socially “undesirable” answers to questions about their health, hygiene, nutrition, or household. In fact, we assume that under-reporting played a bigger role than over-reporting in our estimates of disease prevalence and food insecurity. In other words, the rates of disease prevalence and food insecurity that we have reported in this paper may be underestimates. Another limitation was that we did not capture other behaviors that could increase risks of diarrhea and STH infection, e.g. not washing one’s hands before eating.

A review by Riley demonstrated that self-report, given by children themselves rather than by parent-proxies, is a reliable and valid measure of children’s health outcomes.^93^ Previous epidemiological studies of school-age children in developing countries used self-report.^94–98^ Previous studies measured the relationship between children’s self-reported gastrointestinal (and respiratory) diseases and school WaSH: Weaver and others,^81^ who measured self-reported diarrhea and vomiting; Otsuka and others,^72^ who measured self-reported diarrhea and respiratory symptoms; Chard and Freeman,^99^ who measured self-reported diarrhea and respiratory symptoms and STH infection (unspecified method of assessment). Previous studies (employing a cross-sectional study design) used children’s self-report to assess knowledge, attitudes, and practices (KAP) about STH infection, prior to implementing a school-based intervention.^44, 45, 100^ Cluster-randomized controlled trials (RCTs) measured the impact of school-based WaSH interventions on children’s self-reported KAP about STH infection and STH infection prevalence.^101–103^

Supported by evidence from the above studies, we considered self-reported diarrhea to be a good alternative to other measures of diarrhea, e.g. a review of medical records or interviews with children’s parents. A review of medical records would not have been practical because more time and resources would have been needed to acquire permission to access confidential information and physically inspect hard copies of records for each individual child. Another reason is, because diarrhea is not considered to be a “serious” illness or reportable disease condition, medical records maintained by schools may not have included this type of information. Diarrhea is not usually diagnosed by a physician via a clinical test, but rather by patient-reported symptoms of having loose/watery bowel movements. Interviews with children’s parents would have also been impractical due to increased time and resource constraints. Another reason is, because school-age children do not typically tell their parents about their bowel movements because they think it is unnecessary and/or embarrassing. Thus, had we relied on data from parents’ interviews, rather than data provided directly by children, the information could have been incomplete or inaccurate. Considering the sensitive nature of the topic, it was best to ask the children themselves. Knowing that the children were mature enough to understand the definition we provided for diarrhea and knowing that the children could independently, confidentially, and anonymously answer our questionnaire, we considered self-report to be the ideal outcome measure for diarrhea.

We acknowledge that there are many disadvantages of using self-reported diarrhea in the last month as an outcome measure. For example, some children, in spite of our explanation about the definition of diarrhea, may not have understood what “diarrhea” means. Some children may have misclassified their diarrhea if they forgot that diarrhea meant having >= 3 loose/watery bowel movements in one day. Some children may not have been able to recognize diarrhea if they did not visually inspect their feces in the toilet. Some children may not have wanted to report their diarrhea because of shame or worry that they or their parents would get in trouble or be reported to school or public health authorities. In contrast, other children may have over-reported diarrhea because they hoped to receive medical advice, medical care, special attention, or resources for their school.

Numerous studies have used self-report to measure diarrhea prevalence,^40–42^ and many of these studies^16, 81^ have used a 7-day recall period rather than one month. We conducted our study only during the end of the dry season. However, diarrhea may be more prevalent during the rainy season in the Philippines. Therefore, we used a one-month recall period to capture more cases of diarrhea, some of which may have been missed if we only asked children about the last 7 days. We expect any inaccuracies from our study to not be differential with respect to exposure to inadequate school WaSH; i.e., we do not anticipate that exposure to school WaSH affected the accuracy of children’s self-reported diarrhea. Thus, we assume that any non-differential misclassification of outcomes would bias our findings toward, not away, from the null. Future studies using recall periods shorter than one-month may show more pronounced diarrhea risk, depending on the season (dry v. wet). However, using clinical specimens to ascertain diarrhea may show decreased diarrhea risk, as more children could feel more embarrassed to provide researchers with specimens of their loose/watery feces. Self-reported diarrhea does not differentiate between the types of infection caused by bacteria, viruses, or protozoa, nor does it account for asymptomatic or subclinical infections. A growing body of evidence suggests that many young children from LMICs experience asymptomatic gut colonization with enteric pathogens.^104, 105^ Future studies using different diarrhea measures that capture subclinical infections and asymptomatic pathogen carriage would enable a more nuanced understanding about the disease risks that inadequate school WaSH pose to schoolchildren.

Evidence about the reliability and validity of using self-report to assess STH infection prevalence is very limited. Our use of this outcome measure is supported by evidence from previous studies that used children’s self-report to assess KAP about STH infection.^44, 45, 100^ Furthermore, cluster-RCTs have measured the impact of school-based WaSH interventions on children’s self-reported KAP about STH infection.^101–103^ Of note, in these cluster-RCTs, STH infection-related KAP was supported by STH infection prevalence estimates that were measured via Kato-Katz technique. One limitation of our study was that we used self-report only and did not include other measures of STH infection. In our study, STH prevalence assessments depended on children’s ability to remember if they ever had worms and if they were willing to report it. We framed our questionnaire to illicit this information by including one question: “Have you ever have ‘worms’?” Children were not asked to look at or count worms or eggs in their feces. The lack of fecal visualization and egg counting limited our ability to assess the intensity of STH infection, though this was not an aim of our study. Another limitation was that we did not try to triangulate the information from different methods or sections of the questionnaire. We did not rely on parents to act as proxies for their children by reporting STH infection, nor did we refer to medical records.

In other studies, the intensity of STH infection was defined by the egg count in feces. For example, the Kato-Katz technique, which involves counting the number of eggs per gram of feces, has been widely used to estimate the intensity of STH infection.^6, 24, 106^ A study by Kaminsky and others^107^ used an alternative method, the egg count per direct smear (EDS), to assess the intensity and disease consequences of whipworm infection, caused by *Trichuris trichiura,* in children hospitalized in Honduras. Thirteen children who experienced high (“heavy”) intensity infection, defined as >= 50 eggs per 2 mg of feces, were found to also have anemia, stunting, and malnutrition. However, it is not yet possible to assign a numerical upper limit for the number of eggs or worms in feces that would indicate disease in children. The reason is because the severity of STH infection varies from child to child and depends on many factors, e.g. nutrition status.

In our study, we did not assess the intensity of STH infection, rather we assessed the history of STH infection and nutrition status, factors that play an important role in STH disease development and infection transmission.^108^ Specifically, we assessed children’s nutrition status via anthropometry and self-reported symptoms of severe hunger. We also assessed families’ food insecurity in a subsample of children. We did not use an egg count-based measure because: 1) our study aimed to estimate disease occurrence, not infection intensity or egg distribution; 2) others consider egg counting to have low analytic sensitivity.^109^ Another reason is because STH infection is endemic in the Philippines and public school children are expected, per DepEd policies,^110, 111^ which are in line with the WHO 2017 guidelines,^112^ to undergo deworming twice per year. Recently dewormed children may have no eggs or worms in their feces. Thus, if had we had relied on egg count to estimate STH infection prevalence, we may have ended up with a lower than expected STH prevalence rate that did not accurately reflect disease occurrence.

Another limitation is the reliability of U_sg_ to assess dehydration. Acute dehydration can be assessed via blood tests (e.g. hematocrit, plasma/serum osmolality, serum sodium, hormones), urine tests (e.g. U_osm_, U_sg_, color), clinical features, and subjective complaints of thirst. The gold standard for assessing dehydration is through blood tests that measure plasma/serum osmolality.^113, 114^ However, this test is invasive, expensive, time-consuming, and not practical for field settings or large-sample studies. An alternative is to assess dehydration through urine, which has numerous advantages: it is noninvasive, more affordable, easier to perform (requiring less time, laboratory equipment and expertise), and feasible for large-sample studies.^58^ Urine-based measures depend on several factors: the last urination event, biological differences between individuals, and differences in hormonal responses to dehydration/ rehydration.^115^ U_osm_ is considered to be the gold standard urine-based measure.^54, 55^ Assessment of U_osm_ requires special laboratory equipment, additional time, and technical skills. These reasons make U_osm_ less practical and/or convenient to use in field settings. An alternative is U_sg_, which is highly correlated with U_osm_ ^54, 55^ and has similar sensitivity and specificity,^113^ and it is quicker, easier, more affordable, and requires less laboratory resources. U_sg_ is a quantitative measure of urine’s ratio of solutes (e.g. electrolytes, nitrogenous chemicals) compared with distilled water, which has a specific gravity of 1.000. The normal range of U_sg_ for newborns is 1.001 - 1.020 and for adults is 1.005-1.030, with higher numbers indicating a greater concentration of solutes and, consequently, decreased hydration (known as “dehydration”).^53^ However, there is no U_sg_ level limit associated with disease, only the state of under-hydration. A disadvantage of using U_sg_ is that measurements depend on the number and size of particles contained in the solution.^58^ For example, urine that contains glucose, proteins, and urea could produce falsely elevated U_sg_ values that inaccurately suggest highly concentrated urine. While a review by Zubac and others^116^ indicated that U_sg_ was a reliable measure of dehydration, others have reported that U_sg_ may not be an accurate test of dehydration in children with gastroenteritis.^117^ Due to mixed evidence, it is recommended to use caution when interpreting urine-based measurements of dehydration.

U_sg_ can be measured via refractometry, hydrometry, and (reagent) urine test strips. Urine test strips are affordable, quick and easy to use, and require no laboratory expertise. Another advantage of using urine test strips is the ability to assess a large number of specimens quickly, without the need to disinfect equipment in between assessments, and no risk of cross-contamination between different individuals’ urine specimens. Urine test strips may be preferred in field settings because they allow for point-of-care (POC) measurement that is done on-the-spot, without the need to transport specimens to an external storage facility or laboratory. Others have reported that variability exists between data obtained by urine test strips and data obtained by refractometry, and that urine test strips are not as reliable as refractometry in assessing dehydration.^118–120^ However, others have reported that urine test strips are an acceptable alternative for refractometry in assessing dehydration.^121–123^ Many factors could affect the interpretation of results: timing of last fluid intake, weather, physical activity, and researchers’ interpretation of color change. A disadvantage of using urine test strips is that the accuracy of measurements may decrease as urine alkalinity increases (pH > 7).^124^

Another way to assess dehydration is to examine clinical features, e.g. dry/chapped lips, dry/sticky mouth, cold skin, dry/sunken eyes. However, the appearance of clinical features may vary from child to child. It is possible that some children in our study had an U_sg_ within normal limits but may have actually been dehydrated. In these cases, because we did not assess for clinical features of dehydration, we would have not been able to accurately identify such children as dehydrated. We did not triangulate our measure of dehydration with medical records, reports by teachers or parents, or children’s subjective report of feeling thirsty. Thus, the prevalence rate of acute dehydration that we reported in this paper could be an underestimation.

While *E. coli* is widely used as an indicator of water fecal contamination, it is a measure of only one pathogen and cannot provide a comprehensive water quality assessment. Future studies could provide more information by assessing other bacteria, e.g. *Salmonella typhi* or *Shigella* spp. (“several species”), as well as viruses like Adenovirus and Rotavirus and protozoa like *Cryptosporidium parvum* and *Giardia intestinalis*. Extensive contamination of water by coliform bacteria and *E. coli*, as measured by manual colony counting performed by a single researcher, may have resulted in exposure misclassification due to inaccurate counts or human error. There was a one-year time lag between our measurement of children’s health outcomes and water quality testing. We cannot assure that the water quality we assessed was the same as when we measured children’s health outcomes. Thus, any associations between children’s exposure to poor water quality and disease outcomes may be limited by confounding due to different data collection periods.

Bias due to confounding likely increased during the data analysis phase of our study. We could not control for known confounders because we did not use a case-control study design. Neither nor could we control for unknown confounders by using randomization. Furthermore, caution is strongly encouraged when interpreting the associations measured by our study because we used some outcome measures that have limited reliability and/or validity. Another risk of bias was the large number of children excluded from data analysis due to missing responses. It is important for research studies to clearly state if missing responses include participants who answered “I don’t know”. Regardless of the way that the group of missing responses is categorized, it is important to show that those who were missing responses for key outcomes were not different from those who were not missing responses. In our study, we used STATA to examine missing data: 1) we generated a new variable (“missing”) to identify which children were missing data on key outcomes, e.g. diarrhea only; 2) we counted how many children had missing data; 3) we used multiple logistic regression to assess if any associations were statistically significant. We found no statistically significant difference between children who were missing data and children who were not missing data for key outcomes. Therefore, we concluded that data were missing at random (MAR), though not missing completely at random (MCAR). MCAR means that all data had an equal chance of being missing and that the reason why data were missing is not due to the data themselves.^125^ In an observational study this may be an unrealistic occurrence. In contrast, MAR means that all data, within groups defined by the observed data, had an equal chance of being missing, and that the reason why data were missing is due to a known characteristic of the data themselves.^125^ In our study, the reasons for missing were mostly known: nonresponse (some children did not answer all of the questions in the questionnaire) or enrollment in a school where the school principal declined our request to inspect the school restrooms. In our study, missing responses did not include children who said that they did not know if they ever had a STH infection.

Because of our cross-sectional study design, we could not describe changes in exposures (i.e. risk factors) or outcomes over time, and our ability to described cause and effect was limited. For example, we measured water quality only once, so, we could not assess seasonality/temporal variability, which could greatly impact enteric disease outcomes. Risks of diarrhea and STH infection may be higher during the wet season, but we collected data mostly during the dry season. It is possible that there were no major pathogens circulating in the environment and no infectious diseases outbreaks during this month-long study window. Also, our study duration was perhaps too short to enable us to capture subtle trends in infection intensity, transmission, and spread. Anthropometry data were collected only once, so, we could not assess how exposures to inadequate school WaSH impacted children’s growth over time.

Our analyses of associations with home-level factors were limited by a small sample size of parents of mainly younger (< 13 years old), female children. Thus, our ability to generalize findings to older and male children may be limited. The study lacked a formal sampling frame for enrolling individuals within school clusters. The aim of the study was to study risk factors for three specific diseases (diarrhea, STH, malnutrition), which is easier and more precise to do if the study population is at high risk. Knowing this, we chose to enroll in our study presumed high-risk schools that had high annual enrollment rates. Because STH infection is endemic in the area, the sample was likely biased toward enrolling STH infected-children. The enrollment of schools was largely performed purposively, because of logistical constraints. For example, we did not attempt to enroll schools that were not easily accessible by public transportation, schools undergoing extensive construction, and schools that were located in neighborhoods where the physical safety of researchers would be threatened. Enrollment of participants within schools was restricted to those present at the time of the survey, which is likely to bias the sample toward those being at school more often, such as children with good academic performance or had “hands-on” parents who played an active role in children’s education/upbringing. Children with poor academic performance may have been underrepresented. Because of the nonrandom enrollment of schools and individuals, our study’s diarrhea, STH infection, and malnutrition prevalence estimates, external validity, and generalizability may be limited. We assessed a large number of risk factors, some of which may be unrelated to each other; it is possible that associations have arisen by chance. We also simultaneously assessed multiple outcomes, so, there could have been a possible multiple comparisons effect. This may occur because as the number of tests increases, so does the tendency of seeing a false-positive.^126^ We did not assess schools’ menstrual hygiene management. During our subsequent intervention study, which took place in the same 15 schools but with a different sample of children, we did implement an intervention to address menstrual hygiene management. We will report findings in a forthcoming paper. In light of our study’s limitations, we acknowledge that the validity of the findings we have reported in this paper may be context-specific, i.e. generalizable to a limited number of populations and setting types such as urban poor schoolchildren from LMICs located in the tropics.

### Strengths

We have no intention to overstate the strengths and generalizability of our study’s findings, as we are aware that interpretations of cause and effect associations may be limited due to the observational and cross-sectional study design, which is inherently susceptible to bias due to confounding. To reduce the risk of confounding due to recall and outcome misclassification biases, we provided children with: a private space and sufficient time to answer questionnaires anonymously, easy-to-understand definitions of diarrhea and STH infection, and reassurance that their questionnaire answers would remain confidential and would not impact their school grades.

The rationale for using self-report to measure primary outcomes, in addition to convenience and affordability, was the ability to collect data quickly, easily, and accurately from a large sample of schoolchildren. It has been demonstrated that self-report can be used to assess school-age children’s health.^93^ In fact, self-report is widely used in epidemiological studies involving schoolchildren from developing countries.^95–99^ We acknowledge that while evidence about the reliability and validity of using self-report to assess health is robust for certain health-related outcomes (e.g. pain, management of cancer symptoms, psychopathology), it is mixed or even weak for other outcomes (e.g. physical activity, sleep quality). For this reason, caution is encouraged when interpreting findings.

We assessed not only students’ experience of diarrhea, but also diarrhea frequency, severity (duration [in days] of illness and related hospitalization; disease-related school absence), and spread (number of people at home who also had diarrhea). Students and parents were blinded to objectively measured WaSH inadequacies and water quality testing results from schools, which minimized bias in assessing the link between WaSH exposures and disease outcomes. We used a consistent reporting period to minimize reporting bias. Furthermore, we measured the impacts of over-crowding in schools on disease prevalence rates, highlighting the need for sex-specific or gender-sensitive interventions.

Our study provided new information about using children’s dehydration, measured via U_sg_, as an indicator of schools’ water insecurity or scarcity, which we assessed by children’s self-report and researchers’ observations. Both types of data are helpful for interpreting study findings about children’s dehydration, which is likely a more accurate indicator of water access and water security in schools. It is important to provide children with sufficient drinking water at school to reduce the risk of dehydration, which could negatively impact children’s cognitive performance, e.g. decreased short-term memory.^56^ Evidence about dehydration’s negative impact on cognitive performance has been mostly limited to studies of adults.^130, 131^ Of the few studies^132–135^ that have examined the relationship in children, none used a biometric measure of dehydration. Findings from our study, which used U_sg_ to measure dehydration and children’s self-report and researchers’ observations to measure water insecurity, contributes new information to better understand this topic.

To reduce bias due to confounding, we controlled for known covariates and adjusted for correlation during data analysis by using multiple logistic regression, clustered by school. This enabled us to estimate aORs as a measure of association between exposures and selected outcomes. Clustered observations are different from independent observations because observations fall into clusters, where observations from different clusters are independent but observations within the same cluster are not independent. In our study, we designated schools as the cluster unit, then grouped children into clusters of schools. Clustered errors are errors that are correlated within a cluster (in our study, a single school) but not correlated across clusters. For example, this could happen when children enrolled in one school are correlated; this is known as “within group correlation” or “intragroup correlation”. Clustering by school enabled us to control for potential within group correlation among children from the same school and enabled us to adjust the standard error of estimates. It is important to be able to control for this type of grouping scheme (clustering by school) especially if the independent variable, e.g. WaSH policy, is a constant (i.e. it does not change) in the school cluster.^136^

### Generalizability

The generalizability, or external validity, of our study was supported by: the use of multistage cluster sampling of students in grades 5, 6, 7, 9, and 10 from 15 public schools in 3 cities of Metro Manila as the basis of our diarrhea, STH infection, and malnutrition prevalence and effect estimations; a high participation rate from large sample of children and adolescents; an analysis of risk factors of diarrhea and STH infection, while taking into account variance in prevalence rates across different schools. We used population-based research methodology involving multi-stage sampling of schools from a variety of urban settings and the replication of a demographically diverse study sample of sufficient power--all of which are ways to support the generalizability of study findings. Adherence to study protocols, rigorous investigation, and standardized data collection and reporting increase our confidence that our study findings may be generalizable to other urban poor populations living in areas with comparable weather and school WaSH conditions. Regions with greater access to handwashing basins at schools or populations with good handwashing practices may have fewer exposures to pathogens that cause diarrhea or STH infection. But because the WaSH conditions of our study schools were similar to those of many public schools in Luzon, Philippines,^76, 137, 138^ where Metro Manila is located, our findings may be relevant to other populations of urban poor schoolchildren in other parts of central/southern Luzon, Philippines. However, due to our study’s design and limited capacity to control for confounding and describe cause and effect, findings may be applicable only to specific locations and must therefore be interpreted with caution. Our findings would need to be verified by those reported by larger-scale studies, especially longitudinal studies and RCTs, before any attempts are made to generalize them to diverse study populations or settings in other LMICs.

### Implications for Policy, Practice, Future Research

Our findings point to a knowledge gap characterizing the relationship between exposures to inadequate WaSH in schools and students’ diarrhea, STH infection, and malnutrition. It is difficult to understand this relationship because disease transmission pathways are multifaceted and management of WaSH in schools is shared across multiple sectors. Another knowledge gap exists between the implementation and enforcement of school-based WaSH policies. We have identified 2 policy and practice areas to close this gap and thereby reduce diarrhea and STH infection in students from similar settings: First, a school administrator may be designated to lead a small team of “super-users” (school personnel) to ensure restrooms are being cleaned and maintained, provide hygiene lessons, and promote group handwashing among students. Second, a group of student volunteers may be named as “restroom monitors” in each school to ensure that WaSH facilities are properly used and kept clean, and to remind their fellow students to wash their hands after using the toilet/urinal. The promotion of proper handwashing is urgently needed, not only to prevent diarrhea and STH infection but also to help children thrive during the “new normal”. Now more than ever, handwashing must be promoted to better protect children’s health in light of the on-going Coronavirus disease 2019 (COVID-19) pandemic and the push to re-open schools and keep them open without endangering human life.

Future research needs to explore other methods of assessing diarrhea and STH infection, although our findings indicate that self-report should not necessarily be dismissed. We identified an association between over-nutrition and diarrhea, challenging the conclusions of others who identified, in contrast, that wasting (low weight-for-height) was a risk factor of diarrhea^1^ or was associated with increased diarrhea severity.^139, 140^ However, the association we found was not significant, so, findings should be interpreted with caution. Unlike some other studies,^141^ we did not identify stunting as a significant independent risk factor of diarrhea. Yet, our finding was consistent with those of Bray et al.,^142^ who also reported no significant association between stunting and increased odds of diarrhea. Future longitudinal studies are needed to better understand the long-term impact of diarrhea on stunting. Contamination of water with coliform and *E. coli* was common in our sample, although we found no significant associations with diarrhea or STH infection. More research is needed to understand the relationship between water quality and diarrhea in the Philippines. Another possibility is to replicate the study in private schools in the Philippines. Interesting future studies could explore school WaSH facilities and students’ diarrhea and STH infection prevalence in other countries in the Global South, e.g. Vietnam and Thailand, where temperatures and rainfall are as high as in the Philippines, but the rates of diarrhea- and STH infection-related mortality are, in contrast with our sample, lower.

Moving forward, a cluster-randomized controlled trial (RCT) would be a valuable next step to provide support the associations identified by our study. Research is needed to test WaSH interventions aimed at preventing diarrhea and STH infection through the improvement of children’s nutrition status. This could be achieved by reducing children’s exposure to enteropathogens in schools’ WaSH facilities and increasing children’s health literacy and promoting effective hygiene behaviors, especially handwashing. Greater emphasis should be placed on improving water quality in settings where water contamination is prevalent and is likely a pre-dominant underlying cause of disease.

## Conclusions

Although inadequate WaSH in schools is widely recognized as a risk factor of diarrhea and STH infection, the specific impacts of insufficient WaSH facilities and water supply, lack of restroom cleanliness, and students’ hygiene practices on disease prevalence in Metro Manila, Philippines, were previously unknown. Findings from our study addressed these knowledge gaps by characterizing, via a mixed-methods approach, the WaSH situation in 15 public schools and the prevalence of diarrhea, STH infection, malnutrition, and dehydration in schoolchildren. By linking schools’ WaSH facilities and students’ handwashing and dissatisfaction with and avoidance of school restrooms with disease prevalence, we provide evidence to support the continued use and fine-tuning of comprehensive school-based WaSH interventions and environmental health and education programs, specifically those which promote proper handwashing. Our findings demonstrate that new school WaSH strategies are needed, on the one hand, to protect children from fecal-contaminated water that drives disease risks, and on the other hand, to ensure children have access to the benefits of water security: good health, hygiene, and hydration. Our study is limited by its cross-sectional, observational design; we make no attempt to measure causal inferences. More studies are needed to understand the complex relationship between schools’ WaSH facilities and schoolchildren’s hygiene practices and diarrhea, STH infection, and malnutrition in the Philippines.

### Data sharing

Additional information may be found in the Supplemental Materials. Examples of our questionnaires, unpublished data, and other data collection tools are available as preprints on the open-access online archive, medRxiv https://www.medrxiv.org/.

## Supporting information

Unpublished supplemental materials

## Data Availability

Data are available upon request made to the corresponding author.

