## Supplemental Materials for "Diarrhea, Helminth Infection, Dehydration, and Malnutrition Associated with Water, Sanitation, and Hygiene Facilities and Poor Handwashing in Schools in Metro Manila, Philippines: A Cross-Sectional Study"

**Table S1.** Questions from children's questionnaire

| Number | Question |
|---|---|
| 1 | ID |
| 2 | What is today's date? |
| 3 | What is your birth date? |
| 4 | What is your telephone / cellphone number? |
| 5 | What is your complete address? |
| 6 | What is your age in years? |
| 7 | What is your grade level? |
| 8 | What is your sex? |
| 9 | What is the name of your school? |
| 10 | Have you been attending this school for at least 6 months? |
| 11 | Have you NOT needed to miss school a lot in the past year because of any health problems? |
| 12 | Are you satisfied ("happy") with the school toilets? |
| 13 | Do you use the school toilets every day if needed? |
| 14 | Do you NOT avoid using the school toilet (and just wait until you go home to go to the bathroom)? |
| 15 | Are toilets easily accessible (easy to get to or not too far away)? |
| 16 | Are the toilet rooms easy to use for persons with disabilities (with ramps/stairs/holder, spacious)? |
| 17 | Are there RARELY long lines to use the school toilet? |
| 18 | Are there enough toilets at your school? |
| 19 | Is toilet paper available all the time? |
| 20 | Are the toilet rooms clean? |
| 21 | Do you think that there is enough privacy in the toilet cabins/in front of the urinals? |
| 22 | Can pupils complain to school staff about a bad situation in the school toilet? |
| 23 | Does the school staff take any action to fix the problem when pupils complain? |
| 24 | Do you wash your hands at school? |
| 25 | Is there a hand wash area in your school? |
| 26 | Are you satisfied ("happy") with the hand wash area at school? |
| 27 | Is there always soap available? |
| 28 | Is there always water available for bathroom use? |
| 29 | Is there a towel or paper to dry your hands? |
| 30 | Are the hand wash areas clean at school? |
| 31 | Do you use soap for washing hands at school? |
| 32 | Do they teach good hygiene practices at school? |
| 33 | Do you eat at least 3 meals every day? |
| 34 | Do you eat lunch at school? |
| 35 | Do you RARELY feel hungry? |
| 36 | Do you RARELY feel so hungry that you cannot concentrate at school? |
| 37 | Do you RARELY feel so hungry that you cannot fall asleep? |
| 38 | Do you feel healthy overall? |
| 39 | Have you NEVER had intestinal worms? |
| 40 | In the last 6 months, were you NEVER hospitalized? |
| 41 | In the last 6 months, have you NOT had surgery? |

42 How many times have you had diarrhea in the last month?
43 How many people in your house also had diarrhea in the last month?
44 How many days have you been hospitalized due to diarrhea?

**Table S2.** Results of water quality testing[a], school and home samples (*N* = 60)

| | Schools | | | Homes | | | Normal range | Ref. |
|---|---|---|---|---|---|---|---|---|
| | valid *n* | *n* | % (95% CI) | valid *n* | *n* | % (95% CI) | | |
| % of schools whose water had coliform bacteria[a] | 11 | 4 | 36.4 (10.9, 69.2) | 7 | 3 | 42.9 (9.9, 81.6) | | |
| % of schools whose water had *E.coli* [b] | 12 | 3 | 25.0 (5.5, 57.2) | 7 | 4 | 57.1 (18.4, 90.1) | | |
| Number of water samples with coliform bacteria | 17 | 7 | 41.2 (18.4, 67.1) | 14 | 6 | 42.9 (17.7, 71.7) | | |
| Number of water samples with *E.coli* | 20 | 3 | 15 (3.2, 37.9) | 14 | 4 | 28.6 (8.4, 58.1) | | |
| Air temperature (°C), median (IQR) | | 32 (30.5, 34.0) | | | 33 (32, 34.5) | | 24, 31 | |
| Water temperature (°C), median (IQR) | | 30 (27.0, 32.5) | | | 28.5 (28, 29.4) | | 26.4, 28.3 | |
| BOD, median (IQR) | | 0.63 (0.31, 1.61) | | | 0.88 (0.52, 1.81) | | < 5 mg/L | EPA[c] |
| DO, median (IQR) | | 7.35 (6.33, 8.37) | | | 7.67 (7.37, 8.50) | | 7.81, 8.09 mg/L | EPA[c] |
| Nitrate[d], median (IQR) | | 0.10 (0.05, 0.13) | | | 0.10 (0.08, 0.11) | | < 50 mg/L | DOH[e] |
| pH, median (IQR) | | 7.45 (7.29, 7.63) | | | 7.38 (7.33, 7.61) | | 6.5, 8.5 | DOH[e] |
| TDS , median (IQR) | | 81 (60, 106) | | | 68 (62, 102) | | < 600 mg/L | DOH[e] |
| Turbidity, median (IQR) | | 0.16 (0.01, 1.19) | | | 0.22 (0.11, 0.32) | | < 5 NTU | DOH[e] |

BOD = biological oxygen demand; °C = degree Celsius; CI = confidence interval; DO = dissolved oxygen; DOH = Department of Health, Philippines; *E.coli* = *Escherichia coli*; EPA = United States Environmental Protection Agency; IQR = interquartile range; NTU = nephelometric turbidity units; TDS = total dissolved solids; WHO = World Health Organization.

[a]We collected water samples from a subset of consenting households, located in or nearby the neighborhood of 11 of the study schools, to characterize the water quality in the community. We conducted water sampling after the intervention phase of our larger research project, "WaSH in Manila Schools". We report the study protocols of the intervention study in a forthcoming paper (Sangalang et al., 2020). We collected water per protocol in clean sample containers with lids. We labeled water samples per protocol and, depending on the water quality test we desired, we stored water samples in insulated cool boxes for transport to the laboratory. We used calibrated digital field meters and specialized laboratory equipment to assess water's physical and chemical properties: BOD, DO, nitrates, pH, temperature, TDS, and turbidity. We analyzed samples, using an Extech Instruments (model DO700) (Extech Instruments Corp., Waltham, MA, U.S.A.) water quality meter, at the Institute of Environmental Science and Meteorology at the University of the Philippines Diliman. Other water samples were prepared for transfer to a nearby DOH-accredited water testing laboratory for analysis. We outsourced these water quality tests because our local laboratory had insufficient equipment, materials, and space for analyses or storage. Results from schools and homes were compared to assess if drinking water was contaminated due to unsafe water storage or hygiene practices. We calculated % by dividing the number affected (*n*) by the total number available (valid *n*). Then we multiplied the answer by 100 to get a proportion.

[b]We assessed water for fecal contamination as evidenced by coliform bacteria and *E. coli*. We used a pipette to transfer 1 mL of water to 3M™ Petrifilm™ E. coli/Coliform Count Plates. We photographed plates after 24 hours and 48 hours, then we manually counted the number of colonies on the plates.

[c]Source: EPA. (2006). Voluntary Estuary Monitoring Manual. Chapter 9: Dissolved Oxygen and Biochemical Oxygen Demand https://www.epa.gov/sites/production/files/2015-09/documents/2009_03_13_estuaries_monitor_chap9.pdf

[d]Nitrate as NO3-.

[e]Source: DOH. (2017). Philippine National Standards for Drinking Water of 2017. Administrative Order 2017-0010. https://dmas.doh.gov.ph:8083/Rest/GetFile?id=337128

**Table S3.** Prevalence of health outcomes--diarrhea, STH infection, stunting, undernutrition, over-nutrition, and acute dehydration as evidenced by highly concentrated urine--by sex and age group ($N$ = 1,558 students)

| Outcome | All ($n$ = 1,558) | | | Male ($n$ = 697) | | | Female ($n$ = 861) | | |
|---|---|---|---|---|---|---|---|---|---|
| | valid *n*[a] | *n* | % (95% CI) | valid *n* | *n* | % (95% CI) | valid *n* | *n* | % (95% CI) |
| Diarrhea | 1478 | 421 | 28.5 (26.2, 30.9) | 657 | 201 | 30.6 (27.1, 34.3) | 821 | 220 | 26.8 (23.8, 30) |
| STH infection | 1483 | 647 | 43.6 (41.1, 46.2) | 659 | 314 | 47.7 (43.8, 51.5) | 824 | 333 | 40.4 (37, 43.9) |
| Stunting[b] | 1497 | 227 | 15.2 (13.4, 17.1) | 667 | 102 | 15.3 (12.6, 18.3) | 830 | 125 | 15.1 (12.7, 17.7) |
| Undernutrition[b] | 1478 | 127 | 8.6 (7.2, 10.1) | 659 | 73 | 11.1 (8.8, 13.7) | 819 | 54 | 6.6 (5, 8.5) |
| Over-nutrition[b] | 1478 | 321 | 21.7 (19.6, 23.9) | 659 | 160 | 24.3 (21.1, 27.7) | 819 | 161 | 19.7 (17, 22.5) |
| sg ≥ 1.020[c] | 1405 | 956 | 68 (65.5, 70.5) | 629 | 432 | 68.7 (64.9, 72.3) | 776 | 524 | 67.5 (64.1, 70.8) |
| sg = 1.020[c] | 1405 | 214 | 15.2 (13.4, 17.2) | 629 | 103 | 16.4 (13.6, 19.5) | 776 | 111 | 14.3 (11.9, 17) |
| sg = 1.025[c] | 1405 | 457 | 32.5 (30.1, 35) | 629 | 193 | 30.7 (27.1, 34.5) | 776 | 264 | 34 (30.7, 37.5) |
| sg = 1.030[c] | 1405 | 285 | 20.3 (18.2, 22.5) | 629 | 136 | 21.6 (18.5, 25) | 776 | 149 | 19.2 (16.5, 22.2) |

| Outcome | Child[d] ($n$ = 1039) | | | Teenager[e] ($n$ = 519) | | |
|---|---|---|---|---|---|---|
| | valid *n* | *n* | % (95% CI) | valid *n* | *n* | % (95% CI) |
| Diarrhea | 998 | 296 | 29.7 (26.8, 32.6) | 480 | 125 | 26 (22.2, 30.2) |
| STH infection | 1003 | 442 | 44.1 (41, 47.2) | 480 | 205 | 42.7 (38.2, 47.3) |
| Stunting | 988 | 136 | 13.6 (11.6, 15.9) | 508 | 91 | 17.9 (14.7, 21.5) |
| Undernutrition | 974 | 70 | 7.2 (5.6, 9) | 503 | 57 | 11.3 (8.7, 14.4) |
| Over-nutrition | 974 | 245 | 25.2 (22.5, 28) | 503 | 76 | 15.1 (12.1, 18.5) |
| sg ≥ 1.020 | 913 | 602 | 65.9 (62.8, 69) | 491 | 354 | 72.1 (67.9, 76) |
| sg = 1.020 | 913 | 147 | 16.1 (13.8, 18.6) | 491 | 67 | 13.6 (10.7, 17) |
| sg = 1.025 | 913 | 303 | 33.2 (30.1, 36.3) | 491 | 154 | 31.4 (27.3, 35.7) |
| sg = 1.030 | 913 | 152 | 16.7 (14.3, 19.2) | 491 | 133 | 27.1 (23.2, 31.3) |

Note: CI = confidence interval; sg = specific gravity (of urine); STH = soil-transmitted helminth; WHO = World Health Organization.
[a]Excludes missing data.
[b]Malnutrition (stunting, undernutrition, and over-nutrition) as defined by the WHO (Table 1).
[c]Acute dehydration as evidenced by highly concentrated urine (sg >= 1.020), which we measured using urine test strips.
[d]Child < 13 years old.
[e]Teenager >= 13 years old.

**Figure S1.** Sex-specific aORs for diarrhea, STH infection, and malnutrition by schools' annual enrollment.

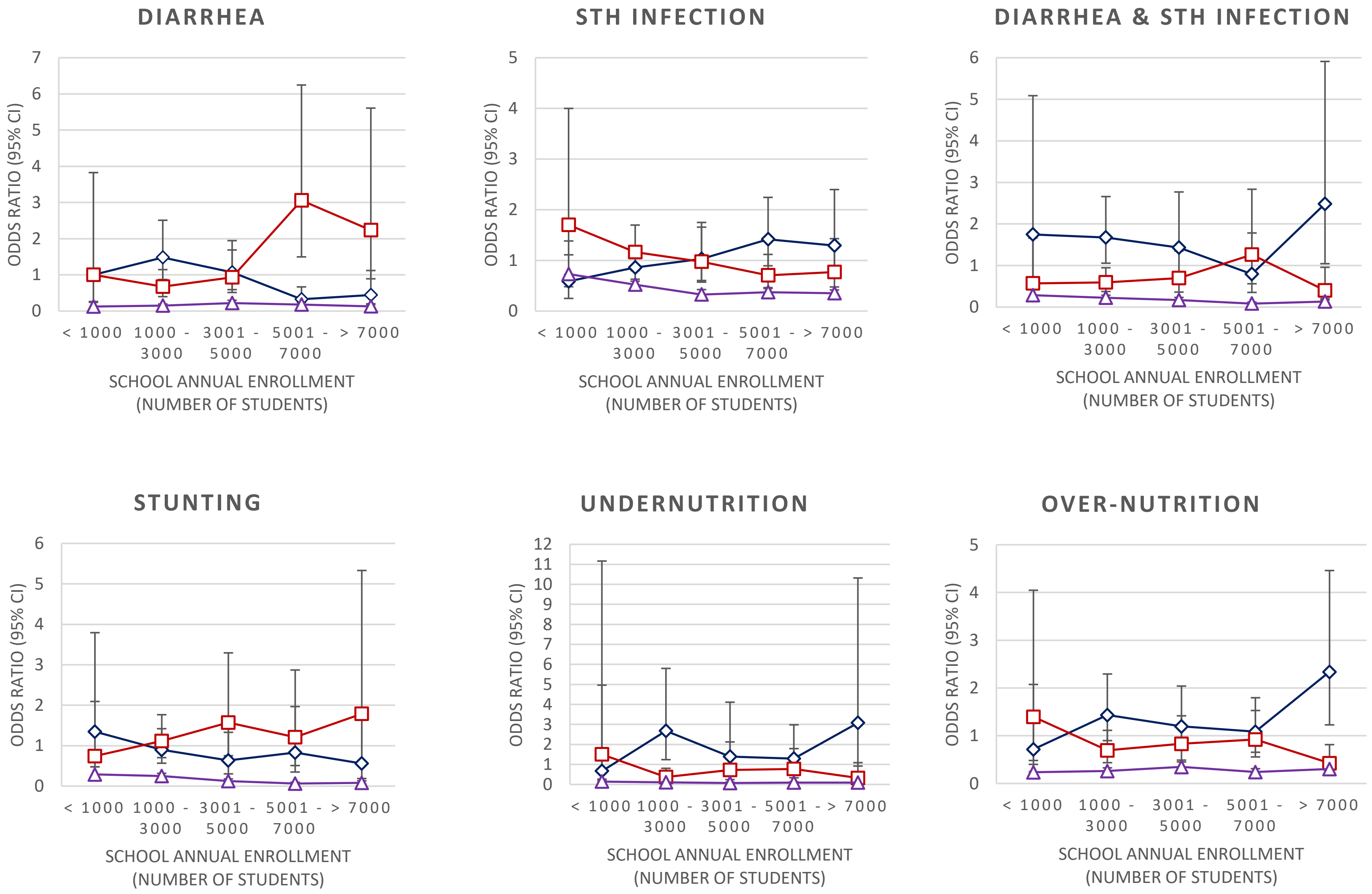

**Figure S2.** Sex-specific aORs for diarrhea, STH infection, and malnutrition by schools' student-to-classroom ratio.

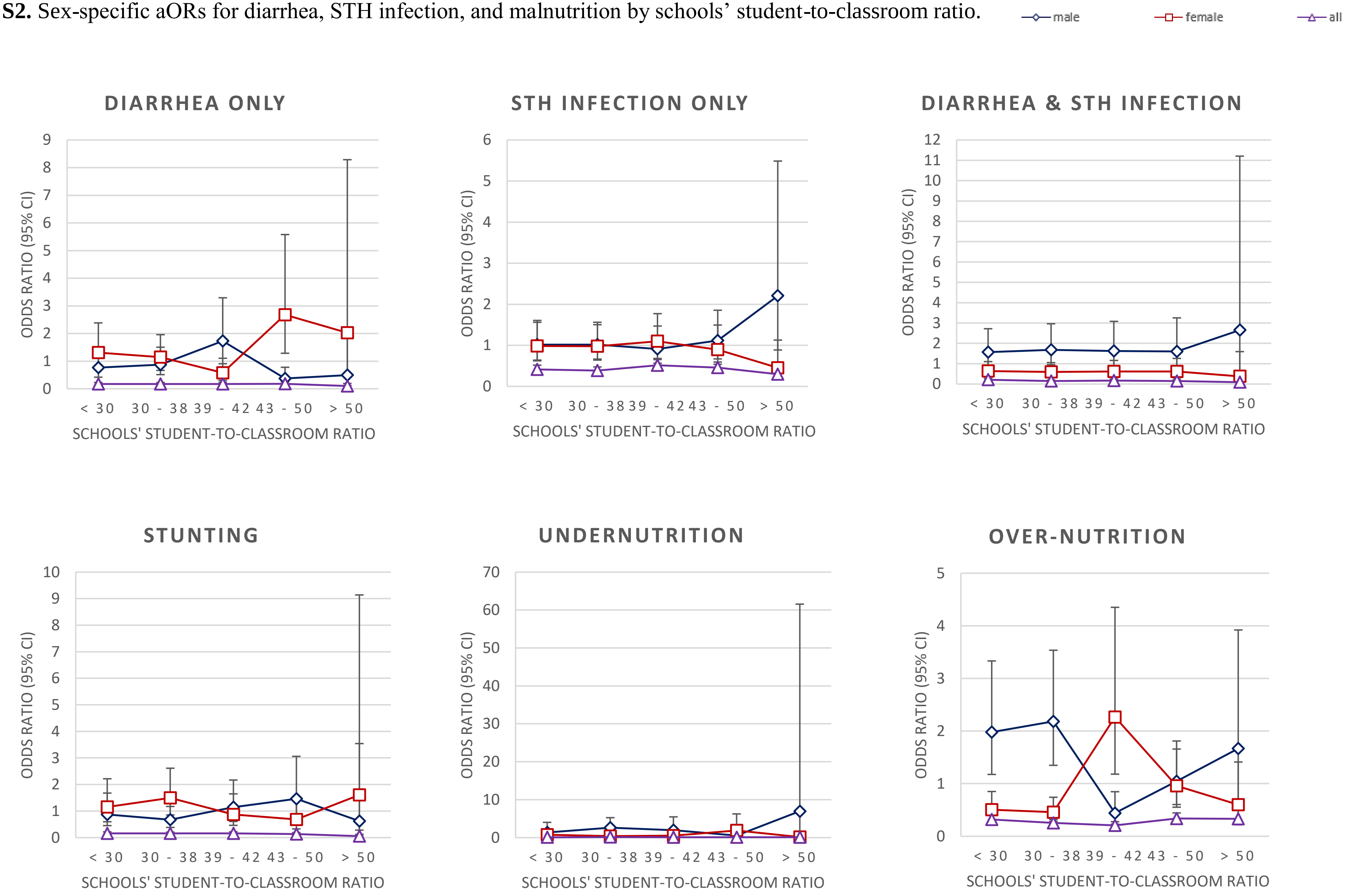

**Box S1.** Equations for logistic regression models

We analyzed the main sample with two logistic regression models: one model (A) for diarrhea only, STH infection only, and dual infection (both diarrhea and STH infection) and one model (B) for malnutrition, i.e. stunting only, underweight only, and over-nutrition only. To measure the association between outcomes and exposures, we used the following model. We list our model's factors in Table S9.

$$\log it(\pi_{abcde}) = \beta_1 San_b \times \beta_2 San_c + Xc_{abc} + \Delta s_b + Ks_d + \zeta_m$$

where

| | |
|---|---|
| $\pi_{abcde}$ | = dichotomous outcome for child *a* in school *b* in cluster *c* in survey *d* in matched group *e* |
| $San_b$ | = whether school has any sanitation |
| $San_c$ | = level of sanitation in school |
| $C_{abc}$ | = vector of child characteristics |
| $S_b$ | = vector of school characteristics |
| $S_d$ | = vector of survey characteristics |
| $\zeta_m$ | = random intercept for matched group *m* that is assumed to be normally distributed with a mean of 0 |

We analyzed the subsample with two logistic regression models: one model (C) for diarrhea only, STH infection only, and dual infection (both diarrhea and STH infection) and one model (D) for malnutrition, i.e. stunting only, underweight only, and over-nutrition only. To measure the association between outcomes and exposures, we used the following model. We list our model's factors in Table S9.

$$\log it(\pi_{abcdefg}) = \beta_1 San_b \times \beta_2 San_c + Xc_{abc} + \Delta s_b + Ks_d + F_f + H_g + \zeta_m$$

where

| | |
|---|---|
| $\pi_{abcde}$ | = dichotomous outcome for child *a* in school *b* in cluster *c* in survey *d* in matched group *e* from family *f* in home *g* |
| $San_b$ | = whether school has any sanitation |
| $San_c$ | = level of sanitation in school |
| $C_{abc}$ | = vector of child characteristics |
| $S_b$ | = vector of school characteristics |
| $S_d$ | = vector of survey characteristics |
| $F_f$ | = vector of family characteristics |
| $H_g$ | = vector of home characteristics |
| $\zeta_m$ | = random intercept for matched group *m* that is assumed to be normally distributed with a mean of 0 |

Note: STH = soil-transmitted helminth.
