## Supplementary material for "Diarrhea, Helminth Infection, Dehydration, and Malnutrition Associated with Water, Sanitation, and Hygiene Facilities and Poor Handwashing in Schools in Metro Manila, Philippines: A Cross-Sectional Study": Unpublished supplemental materials

### Unpublished Supplemental Material

### Table of Contents

**Table S1.** Questions from children's health examination.
**Table S2.** Questions from school principal's interview.
**Table S3.** Questions from school restroom inspection.
**Table S4.** Questions from parent's interview.
**Table S5.** Questions from home restroom inspection.
**Table S6.** Expanded table of main sample descriptives of demographic, health, and hygiene-related factors of students from main sample and WaSH-related structural factors of schools.
**Table S7.** Prevalence rates of health outcomes--diarrhea only, STH infection only, diarrhea and STH infection, stunting only, undernutrition only, and over-nutrition only--by school.

**Table S1.** Questions from children's health examination

| Number | Question |
|---|---|
| 1 | ID |
| 2 | Date of birth |
| 3 | Telephone number |
| 4 | Height (m) |
| 5 | Weight (kg) |
| 6 | Urine pH |
| 7 | Urine Specific Gravity |
| 8 | Urine Glucose |
| 9 | Urine Protein |
| 10 | Do you have any questions/comments? |
| 11 | Research ID |

**Table S2.** Questions from school principal's interview

| Number | Question |
|---|---|
| 1 | What is today's date? |
| 2 | What is the name of your school? |
| 3 | What is your name? |
| 4 | What is your telephone / cellphone number? |
| 5 | What is your email address? |
| 6 | How many students are enrolled in the school? |
| 7 | How many male students are enrolled? |
| 8 | How many female students are enrolled? |
| 9 | How many students have a physical handicap (e.g. cannot walk or is blind)? |
| 10 | Which grade levels are taught at the school (e.g. grades 1-6 or grades 9-12)? |
| 11 | Is there a school policy to clean the students' restrooms at least once per day? |
| 12 | Is there someone who is assigned to cleaning the students' restrooms? (e.g. janitor or cleaning lady) |
| 13 | Is there someone who is assigned to making sure that the cleaning policy is enforced? (e.g. sanitation officer or administrative staff) |
| 14 | Is there a school policy for repairing broken parts in the students' restrooms? |
| 15 | Is there someone assigned to checking if there are broken parts in the students' restrooms at least once per month? |
| 16 | Is hygiene taught in the school curriculum? |
| 17 | Is hand washing taught in the school curriculum? |
| 18 | Are students allowed to report any issues/concerns/problems with the students' restrooms? |
| 19 | Is there someone assigned to address students' issues/concerns/problems with the students' restrooms? |
| 20 | Are you satisfied with the sanitation in the students' restrooms? |
| 21 | Are you satisfied with the hygiene behaviors of the students? |
| 22 | Do you have any other questions or comments? |

**Table S3.** Questions from school restroom inspection

| Number | Question |
|---|---|
| 1 | What is the name of the school? |
| 2 | What is the date of inspection? |
| 3 | Restroom code |
| 4 | Is this restroom for males only? |
| 5 | Is this restroom NOT coed (for both males and females)? |
| 6 | Is the restroom clean? |
| 7 | Is the restroom without a bad smell? |
| 8 | Is the restroom well-lit (that is, not too dark)? |
| 9 | Is the restroom light working? |
| 10 | Is there place to wash hands in the same room? |
| 11 | What type of handwashing facilities are there in/ near this restroom? |
| 12 | Is there water to wash hands? |
| 13 | Is there soap to wash hands? |
| 14 | Is there a towel or paper to dry hands? |
| 15 | Is there toilet paper? |
| 16 | Is the floor dry? |
| 17 | Is there a door that locks for each toilet cabin? |
| 18 | Is there a wall separation for privacy of each toilet cabin? |
| 19 | Is the wall or separation high enough to provide privacy? |
| 20 | For male restrooms, are there partitions between urinals? |
| 21 | Are there NO signs of graffiti? |
| 22 | Are there NO signs of damage (e.g. leaky pipes, cracked walls, open ceilings, peeling paint, flaking plaster)? |
| 23 | Are there NO signs of mold? |
| 24 | Type of toilet |
| 25 | If it is a flush or pour-flush toilet, then where does the wastewater flow to? (see attached guide) |
| 26 | Do ALL of the toilets flush? (if a toilet is marked as “out of order,” then no need to check) |
| 27 | If it is a dry toilet, then what type of toilet is it? (see attached guide) |
| 28 | Are there NO flies? |
| 29 | Is there a garbage can inside? |
| 30 | Number of toilet bowls |
| 31 | Number of urinals |
| 32 | Number of handwashing basins |
| 33 | Distance between two (2) urinals (m) |
| 34 | Distance from toilet/urinal and nearest handwashing basin (m) |
| 35 | If there is a urinal wall present, then how long (cm) is it? Write "N/A" if there is no urinal wall. |
| 36 | Length of the urinal partition (cm) |
| 37 | Width of the urinal partition (cm) |
| 38 | How many photos did you take? |
| 39 | Did you collect a water specimen? |

40 Did you notice anything else that you wish to document?

**Table S4.** Questions from parent's interview

| Number | Question |
|---|---|
| 1 | What is today's date? |
| 2 | What is the name of the student (First and Last Name)? |
| 3 | What is the date of birth of the student? |
| 4 | What is the name of your child's school? |
| 5 | What is the name of the parent (First and Last Name)? |
| 6 | How many people live in the home? |
| 7 | How many adults live in the home? |
| 8 | How many children live in the home? |
| 9 | How long (years) have you lived in the home? |
| 10 | Is their enough food to eat for everyone in your home for most days? |
| 11 | Are there enough places near your home where you can buy food? |
| 12 | Are the prices of the food affordable? |
| 13 | Have you NEVER experienced a time when you wanted to eat but had no food or could not get to buy food? |
| 14 | Have you and your family NEVER had to ask for food? |
| 15 | Do you RARELY feel extreme hunger? |
| 16 | Do you eat a variety of food (e.g. fruits, vegetables, meat, rice) on most days? |
| 17 | Do you cook more often than buy already prepared food? |
| 18 | Do you have water that is safe for drinking? |
| 19 | Do you have enough water so everyone can drink? |
| 20 | Does the person who cooks for the family wash his/her hands before cooking? |
| 21 | Does he/she wash the fruits/vegetables before cooking them? |
| 22 | Do you have any other questions or comments? |

**Table S5.** Questions from home restroom inspection

| Number | Question |
|---|---|
| 1 | Date of inspection |
| 2 | Home address |
| 3 | Student's name |
| 4 | Student's date of birth |
| 5 | Student ID |
| 6 | Name of School |
| 7 | Is the restroom inside the house? |
| 8 | Is the restroom clean? |
| 9 | Is the restroom without a bad smell? |
| 10 | Is there a restroom light? |
| 11 | Is the restroom light working? (e.g., light bulb, bottle light, etc.) |
| 12 | Is the restroom well-lit (that is, not too dark)? |
| 13 | Is there a place to wash the hands in the same room? |
| 14 | Is there water to wash the hands? |
| 15 | Is there soap to wash the hands? |
| 16 | Is there towel or paper to dry the hands? |
| 17 | Is there toilet paper? |
| 18 | Is the floor dry? |
| 19 | Is there a door for the toilet that locks? |
| 20 | Is the toilet an example of "improved sanitation"? (e.g. flush toilet instead of open pit) |
| 21 | Is it a flush toilet? |
| 22 | Does the toilet flush? |
| 23 | Is there a septic tank for the toilet? |
| 24 | Are there NO signs of damage? (e.g. leaky pipes, cracked walls) |
| 25 | Are there NO signs of mold? |
| 26 | Is there a garbage can nearby? |
| 27 | Number of toilet bowls |
| 28 | Number of handwashing basins |
| 29 | Distance from toilet to nearest handwashing basin (in meters) |

**Table S6.** Expanded table of main sample descriptives of demographic, health, and hygiene-related factors of students (*N* = 1,558) from main sample and WaSH-related structural factors of schools (*N* = 15)

| | *n* | % (95% CI) |
|---|---|---|
| Student factors (*N*=1,588) | | |
| WaSH-related knowledge (self-reported) | | |
| Students may not complain about the “bad” condition of school restroom | 312 | 21.0 (18.9, 23.1) |
| Does not know if students may complain or not | 626 | 42.1 (39.6, 44.6) |
| School staff does not respond to students’ complaints about school restroom | 280 | 18.8 (16.8, 20.8) |
| Does not know if school staff responds to complaints or not | 589 | 39.6 (37.1, 42.1) |
| Health history and nutrition (self-reported) | | |
| Has been hospitalized (ever) | 211 | 14.2 (12.4, 16.0) |
| Has had surgery (ever) | 72 | 4.8 (3.7, 5.9) |
| Eats 3 meals per day | 1,394 | 94 (92.7, 95.2) |
| It is “rare” to feel extremely hungry | 1,162 | 78.4 (76.2, 80.4) |
| Experienced being too hungry to focus at school | 242 | 16.3 (14.4, 18.2) |
| Experienced being too hungry to sleep | 199 | 13.4 (11.7, 15.1) |
| Health outcomes | | |
| Had illness (self-reported) | | |
| No diarrhea and no STH infection | 617 | 41.9 (39.3, 44.4) |
| Diarrhea but no STH infection | 212 | 14.4 (12.6, 16.3) |
| STH infection but no diarrhea | 438 | 29.7 (27.4, 32.1) |
| Both diarrhea and STH infection | 207 | 14.0 (12.3, 15.9) |
| Had diarrhea 1 - 3 days last month | 374 | 25.3 (23.1, 27.6) |
| Hospitalized for diarrhea 1 - 3 days last month | 91 | 6.1 (5.0, 7.5) |
| Malnutrition[a] and acute dehydration[b] (observed) | | |
| No stunting and no undernutrition | 1165 | 78.8 (76.6, 80.9) |
| Stunting but no undernutrition | 186 | 12.6 (10.9, 14.4) |
| Undernutrition but no stunting | 87 | 5.9 (4.7, 7.2) |
| Both stunting and undernutrition | 40 | 2.7 (1.9, 3.7) |
| No stunting and no over-nutrition | 946 | 64.0 (61.5, 66.5) |
| Stunting but no over-nutrition | 211 | 14.3 (12.5, 16.2) |
| Over-nutrition but no stunting | 306 | 20.7 (18.7, 22.9) |
| Both stunting and over-nutrition | 15 | 1.0 (1.0, 1.7) |
| Normal weight | 1030 | 69.7 (67.3, 72.0) |
| Urine specific gravity (sg), median (IQR) | | 1.025 (1.015, 1.025) |
| Urine has (any) protein[c] | 360 | 25.7 (23.4, 28.1) |
| Urine protein, trace | 346 | 24.7 (22.5, 27.0) |
| Urine protein, level 1 | 12 | 0.9 (0.4, 1.5) |
| Urine protein, level 2 | 1 | 0.1 (0.00181, 0.4) |
| Urine protein, level 3 | 1 | 0.1 (0.00181, 0.4) |
| Urine has no protein | 1041 | 74.3 (71.9, 76.6) |
| School factors (*N* = 15) | | |

| | | |
|---|---|---|
| Facilities (observed) | | |
| Toilets (*n* = 223) | | |
| Number of male-only toilets, median (IQR) | | 7 (3, 11) |
| Number of female-only toilets, median (IQR) | | 10 (3, 16) |
| Number of coed toilets, median (IQR) | | 2 (2, 5) |
| Male student-to-male toilet ratio, median (IQR) | | 320.4 (245.1, 401.5) |
| Female student-to-female toilet ratio, median (IQR) | | 261.2 (182.1, 364.7) |
| Number of schools that exceeded male student-to male toilet ratio | 11 | 91.7 (61.5, 99.8) |
| Number of schools that exceeded female student-to female toilet ratio | 12 | 92.3 (64, 99.8) |
| Urinals (*n* = 49.5[d]) | | |
| Number of urinals, median (IQR) | | 2 (0, 4.9) |
| Male student-to-urinal ratio, median (IQR) | | 608.6 (407.8, 1624.5) |
| Number of schools that exceeded male student-to urinal ratio | 8 | 88.9 (51.8, 99.7) |
| Handwashing basins (*n* = 129) | | |
| Number of handwashing basins not near toilet | 56 | 45.1 (36.2, 54.3) |
| Mean distance (m) between handwashing basin and toilet, median (IQR) | | 1.8 (0.9, 2.9) |
| Policies and school principals' perceptions (self-reported) | | |
| School has a policy to clean school restroom daily | 14 | 93.3 (68.1, 99.8) |
| School has a policy to enforce daily cleaning of school restroom | 14 | 93.3 (68.1, 99.8) |
| School principal is not satisfied with students' hygiene | 7 | 46.7 (21.3, 73.4) |
| School principal is not satisfied with restrooms' sanitation | 7 | 46.7 (21.3, 73.4) |
| Students' perceptions (self-reported) | | |
| Is not satisfied with restroom | 716 | 51.8 (49.2, 54.3) |
| Restroom lacks cleanliness | 924 | 62.1 (59.7, 64.6) |
| Restroom provides insufficient privacy | 808 | 54.4 (51.9, 57.0) |
| Restroom is too far away | 114 | 8.2 (6.7, 9.6) |
| Restroom is not accessible for PWD | 777 | 52.2 (49.7, 54.8) |
| Restroom has long line to use toilet | 351 | 23.6 (21.4, 25.8) |
| Number of school restrooms is not sufficient for the number of users | 495 | 33.3 (30.9, 35.7) |
| No toilet paper available | 1302 | 87.5 (85.8, 89.2) |
| Is not satisfied with handwashing basin | 259 | 23.8 (21.3, 26.4) |
| Handwashing basin lacks cleanliness | 335 | 30.8 (28.1, 33.6) |
| No water available | 241 | 22.1 (19.7, 24.6) |
| No soap available | 848 | 78.1 (75.6, 80.5) |
| No towel or paper for drying hands | 946 | 87.0 (85.0, 89.0) |
| No handwashing basin available | 398 | 28.6 (26.2, 30.9) |

Note: CI = confidence interval; IQR = interquartile range; PWD = person(s) with disabilities; sg = specific gravity; STH = soil-transmitted helminth; WHO = World Health Organization.

[a]Malnutrition as defined by the WHO (Table 1).

[b]Acute dehydration, as exhibited by highly concentrated urine, was defined as urine sg >= 1.020.

[c]We measured urine protein by using urine test strips (Insight Urinalysis Reagent Strips, Acon Laboratories Inc., San Diego, California, U.S.A.). The urine protein values are as follows: trace/+1 block (20 mg/dL), level 1/+2 blocks (50 mg/dL), level 2/+3 blocks (100 mg/dL), and level 3/+4 blocks (200, 400 mg/dL).
[d]Urinal $N$ = 49.5 includes data from one school wherein no urinals were present in one of the restrooms for male students. However, inside the restroom there was a common area, with running water, where male students could urinate.

**Table S7.** Prevalence rates of health outcomes--diarrhea only, STH infection only, diarrhea and STH infection, stunting only, undernutrition only, and over-nutrition only--by school (*N* = 1,558)

| School | City | Students Surveyed | Diarrhea | | | STH infection | | | Stunting | | |
|---|---|---|---|---|---|---|---|---|---|---|---|
| | | | valid *n*[a] | *n* | % (95% CI) | valid *n* | *n* | % (95% CI) | valid *n* | *n* | % (95% CI) |
| 1 | Manila | 111 | 111 | 40 | 36 (27.1, 45.7) | 110 | 37 | 33.6 (24.9, 43.3) | 119 | 16 | 13.4 (7.9, 20.9) |
| 2 | Navotas | 108 | 106 | 36 | 34 (25, 43.8) | 106 | 40 | 37.7 (28.5, 47.7) | 108 | 17 | 15.7 (9.4, 24) |
| 3 | Navotas | 90 | 90 | 30 | 33.3 (23.7, 44.1) | 90 | 58 | 64.4 (53.7, 74.3) | 89 | 27 | 30.3 (21, 41) |
| 4 | Navotas | 105 | 103 | 37 | 35.9 (26.7, 46) | 103 | 52 | 50.5 (40.5, 60.5) | 105 | 16 | 15.2 (9, 23.6) |
| 5 | Navotas | 103 | 80 | 34 | 42.5 (31.5, 54.1) | 80 | 55 | 68.8 (57.4, 78.7) | 103 | 24 | 23.3 (15.5, 32.7) |
| 6 | Navotas | 99 | 97 | 31 | 32 (22.9, 42.2) | 97 | 55 | 56.7 (46.3, 66.7) | 98 | 22 | 22.4 (14.6, 32) |
| 7 | Navotas | 133 | 120 | 34 | 28.3 (20.5, 37.3) | 119 | 63 | 52.9 (43.6, 62.2) | 132 | 38 | 28.8 (21.2, 37.3) |
| 8 | Quezon City | 116 | 109 | 19 | 17.4 (10.8, 25.9) | 109 | 34 | 31.2 (22.7, 40.8) | 116 | 8 | 6.9 (3, 13.1) |
| 9 | Quezon City | 103 | 103 | 31 | 30.1 (21.5, 40) | 103 | 46 | 44.7 (34.9, 54.8) | 103 | 13 | 12.6 (6.9, 20.6) |
| 10 | Quezon City | 100 | 98 | 16 | 16.3 (9.6, 25.2) | 97 | 28 | 28.9 (20.1, 39) | 100 | 5 | 5 (1.6, 11.3) |
| 11 | Quezon City | 118 | 104 | 21 | 20.2 (13, 29.2) | 103 | 41 | 39.8 (30.3, 49.9) | 118 | 15 | 12.7 (7.3, 20.1) |
| 12 | Quezon City | 106 | 106 | 29 | 27.4 (19.1, 36.9) | 106 | 40 | 37.7 (28.5, 47.7) | 106 | 11 | 10.4 (5.3, 17.8) |
| 13 | Quezon City | 90 | 86 | 24 | 27.9 (18.8, 38.6) | 86 | 27 | 31.4 (21.8, 42.3) | 90 | 5 | 5.6 (1.8, 12.5) |
| 14 | Quezon City | 82 | 80 | 19 | 23.8 (14.9, 34.6) | 80 | 25 | 31.3 (21.3, 42.6) | 82 | 7 | 8.5 (3.5, 16.8) |
| 15 | Quezon City | 94 | 85 | 20 | 23.5 (15, 34) | 94 | 46 | 48.9 (38.5, 59.5) | 36 | 3 | 8.3 (1.8, 22.5) |
| Total | | 1558 | 1478 | 421 | | 1483 | 647 | | 1505 | 227 | |

Note: CI = confidence interval; STH = soil-transmitted helminth.
[a]Excludes missing data.

| School | City | Students Surveyed | Undernutrition | | | Over-nutrition | | |
|---|---|---|---|---|---|---|---|---|
| | | | valid *n* | *n* | % (95% CI) | valid *n* | *n* | % (95% CI) |
| 1 | Manila | 111 | 111 | 7 | 6.3 (2.6, 12.6) | 111 | 35 | 31.5 (23, 41) |
| 2 | Navotas | 108 | 108 | 4 | 3.7 (1, 9.2) | 108 | 32 | 29.6 (21.2, 39.2) |
| 3 | Navotas | 90 | 89 | 11 | 12.4 (6.3, 21) | 89 | 17 | 19.1 (11.5, 28.8) |
| 4 | Navotas | 105 | 105 | 6 | 5.7 (2.1, 12) | 105 | 16 | 15.2 (9, 23.6) |
| 5 | Navotas | 103 | 103 | 10 | 9.7 (4.8, 17.1) | 103 | 21 | 20.4 (13.1, 29.5) |
| 6 | Navotas | 99 | 98 | 6 | 6.1 (2.3, 12.9) | 98 | 20 | 20.4 (12.9, 29.7) |
| 7 | Navotas | 133 | 132 | 20 | 15.2 (9.5, 22.4) | 132 | 16 | 12.1 (7.1, 18.9) |
| 8 | Quezon City | 116 | 116 | 8 | 6.9 (3, 13.1) | 116 | 29 | 25 (17.4, 33.9) |
| 9 | Quezon City | 103 | 103 | 10 | 9.7 (4.8, 17.1) | 103 | 22 | 21.4 (13.9, 30.5) |
| 10 | Quezon City | 100 | 100 | 12 | 12 (6.4, 20) | 100 | 17 | 17 (10.2, 25.8) |
| 11 | Quezon City | 118 | 118 | 13 | 11 (6, 18.1) | 118 | 19 | 16.1 (10, 24) |
| 12 | Quezon City | 106 | 106 | 8 | 7.5 (3.3, 14.3) | 106 | 27 | 25.5 (17.5, 34.9) |
| 13 | Quezon City | 90 | 90 | 2 | 2.2 (0.2, 7.8) | 90 | 18 | 20 (12.3, 29.8) |
| 14 | Quezon City | 82 | 82 | 7 | 8.5 (3.5, 16.8) | 82 | 26 | 31.7 (21.9, 42.9) |
| 15 | Quezon City | 94 | 17 | 3 | 17.6 (3.8, 43.4) | 17 | 6 | 35.3 (14.2, 61.7) |
| | Total | 1558 | 1478 | 127 | | 1478 | 321 | |

Note: CI = confidence interval; STH = soil-transmitted helminth.
[a]Excludes missing data.
